# Demographic Factors Moderate the Effectiveness of Obesity Prevention Interventions: A Secondary Analysis of College Intervention Trials

**DOI:** 10.64898/2026.04.22.26351238

**Authors:** Caitlyn Winn, Leah Groene, Sarah Colby, Lilian Ademu, Melissa D. Olfert, Carol Byrd-Bredbenner, Anne Mathews, Jesse Stabile Morrell, Priscilla Brenes, Onikia Brown, Makenzie Barr-Porter, Geoffrey Greene, Jaapna Dhillon

## Abstract

**Background:** College-attending young adults frequently experience declines in diet quality, physical activity, and psychological well-being during the transition to independent living, contributing to weight gain during the first year of college. Although multicomponent lifestyle interventions have been developed to address these behaviors, the responsiveness to such programs could differ across demographic factors associated with health behaviors, such as sex, race, and ethnicity. Hence, this secondary analysis of large-scale college health trials evaluated whether the effectiveness of such interventions differed by these demographic factors.

**Methods:** Data were combined from two multi-site randomized controlled trials: Young Adults Eating and Active for Health (YEAH) trial and the Get FRUVED trial. Both interventions used theory-based approaches to promote healthy weight management through improvements in diet quality, physical activity, and stress management. Baseline-adjusted linear regression models evaluated the effects of group (intervention, control) and its interactions with sex, race (White, Black, Other), or Hispanic ethnicity. Models were adjusted for baseline outcome values, baseline BMI, study (YEAH vs. FRUVED), and state of data collection.

**Results:** Intervention participants reported higher fruit and vegetable intake, lower processed meat intake, and longer sleep duration compared with controls. However, there was significant heterogeneity in these dietary outcomes by ethnicity, race, and sex. Non-Hispanic participants in the intervention group had higher fruit and vegetable intake compared to controls (p < 0.05). And, within the intervention group, Hispanic females had lower bacon/sausage intake than Hispanic males and non-Hispanic females (p < 0.05). With respect to race, Black participants reported higher total processed meat intake than White and Other race participants in the intervention group (p <0.05). These demographic factors did not moderate the intervention’s impact on physical activity, sleep duration, and perceived stress. Overall, the intervention appeared to be the least effective for Hispanic males who exhibited higher body weight and waist circumference compared with Hispanic females and non-Hispanic males (p < 0.05).

**Conclusions:** Multicomponent lifestyle interventions can improve selected dietary outcomes among college students, but effectiveness may differ across demographic subgroups. Culturally and sex-tailored strategies that consider the intersecting influences of sex, race, and ethnicity may enhance intervention effectiveness during the transition to college.

## INTRODUCTION

College students, typically ages 18-25, face several health challenges as they transition to independent living. Poor diet quality, inadequate physical activity, and high levels of academic stress are consistently reported among this population (1–4). As a result, a significant proportion of students experience significant weight gain during their first year of college, averaging 1.6-3.5 kg (3.5-7.7 lb), with this upward trend continuing throughout their college years (5–9). Overall, more than one-third of first-year college students’ body mass index (BMI) fall in the categories of overweight (30.5%) or obese (8.6%) (4). Low intake of fruits, vegetables, and low-fat dairy products and high consumption of ultra-processed foods (UPFs) are characteristics of this cohort (1,2) and may play a significant role in observed weight trends. Notably, adolescents and young adults consume more UPFs than any other age group averaging 56.1–67% of their total energy intake from these foods (10). In addition to excessive intake of calorically dense and micronutrient poor UPFs, only 19% of students report regularly consuming a balanced diet (2).

With young adulthood recognized as a formative period for long-term habit development, the suboptimal dietary patterns of college-attending young adults are a major concern. Physical inactivity is also a common trait of this cohort, with more than half (54%) of college students reporting being active less than twice a week, greatly increasing the risk for development of chronic diseases such as type 2 diabetes and heart disease later in life (2,4). Furthermore, first-year college students are particularly vulnerable to mental health challenges; 79% report feelings of sadness or depression, and more than 40% report facing anxiety as they adjust to increased independence and new environments (2,11). Academic stress has also been shown to significantly impact the psychological well-being of college students (12,13).

Given these concerns, interventions promoting healthy lifestyle practices should be encouraged among college students during their first year, when they are particularly vulnerable to behavioral and environmental factors related to increased health risks. Several large-scale, multi-component interventions have been developed to address lifestyle behaviors in college populations (14–16), including the Young Adults Eating and Active for Health (YEAH) trial, which demonstrated overall improvements in diet quality, sleep, and perceived stress among college students (17). The success of project YEAH inspired the development of Fruit and Vegetable Education (FRUVED), a related college health intervention targeting unhealthy weight gain among first-year college students. However, few studies have explored how factors such as sex, race, and ethnicity influence responses to these programs, despite clear evidence that these factors shape health behaviors. For instance, women are more likely to experience stress-related eating, while men tend to engage in higher levels of physical activity (1). In NHANES-based analyses, race/ethnicity differences in adherence to food-based guidelines are observed, with non-Hispanic Black adults often showing lower adherence for several food groups versus non-Hispanic White adults, and Mexican American adults showing distinct patterns in certain foods (e.g., beans) compared with White and Black groups (18). Although studies highlight consistent sex differences and racial and ethnic disparities in diet quality among U.S. adults (18–20), relatively few randomized controlled interventions have evaluated whether the effectiveness of lifestyle programs differs by these factors (21). Even fewer studies have examined whether such disparities influence intervention responsiveness among younger adults during the college transition period, a developmental stage associated with heightened vulnerability to weight gain and lifestyle changes (22). This represents a critical gap given that sociodemographic characteristics influence baseline dietary habits, physical activity, sleep behaviors and perceived stress and could thereby affect intervention responsiveness among college students. Hence, in this secondary analysis of two large-scale, multicomponent, multi-site college health trials we hypothesized that intervention responsiveness among first-year college students would be moderated by the intersecting influences of sex, race, and ethnicity.

## METHODS

### Parent Studies

This study includes a secondary analysis of two multi-site clinical trials: YEAH and FRUVED. Both studies applied community-based participatory research (CBPR) and theory-based approaches to promote healthy weight management in college students by targeting diet quality, physical activity, and stress management. The study protocols were approved by the Institutional Review Boards of the individual sites, and all participants provided written informed consent. FRUVED is registered on Clinical Trials.gov: NCT02941497. The participant flow throughout the studies is depicted in **Figure 1**.

**Figure 1.**
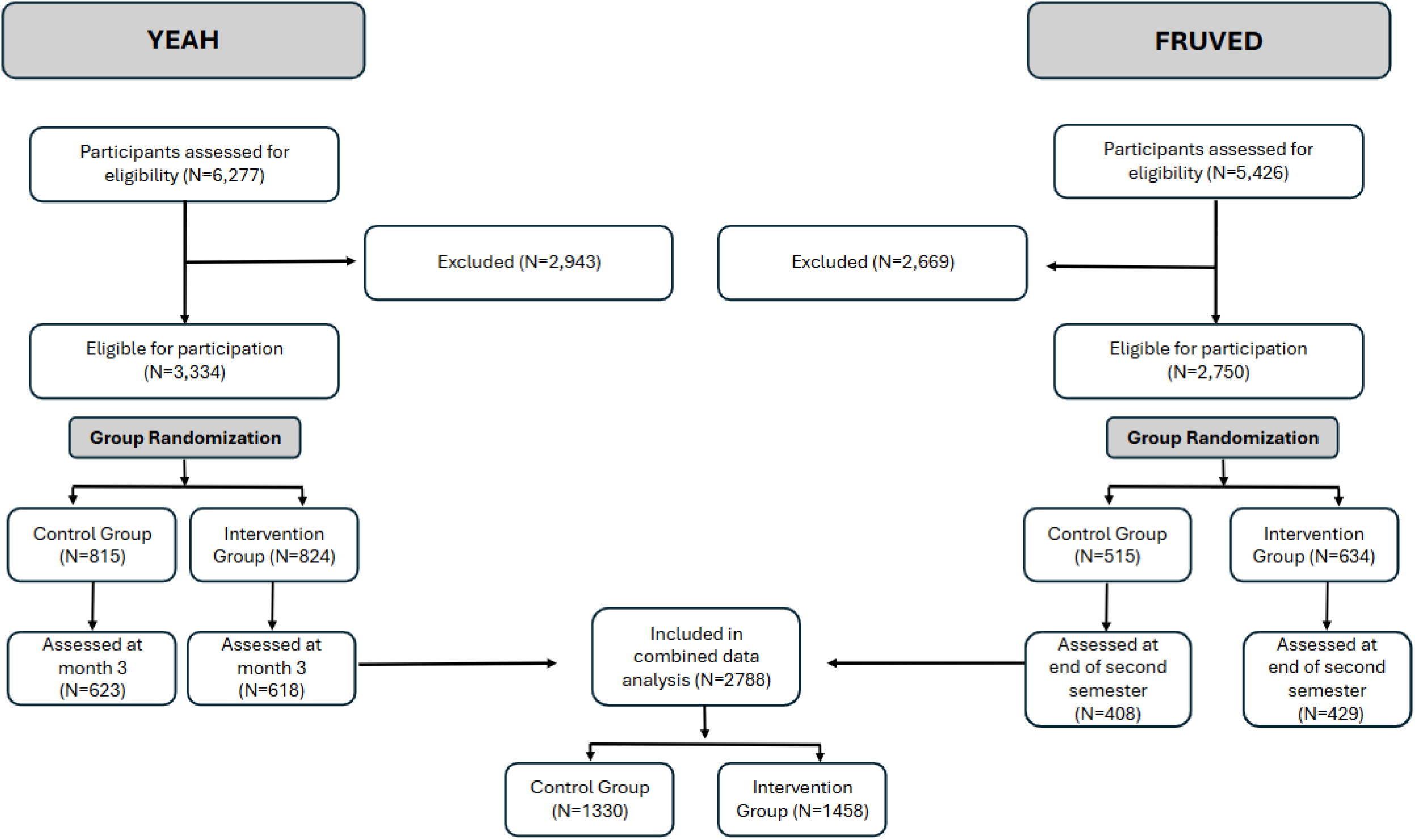
Participant Flow in the Yeah and Fruved Studies.

### YEAH Trial

YEAH was a 15-month web-based randomized controlled trial conducted in 2011 across 13 universities (17,23). Participating universities served as both control and intervention sites and included East Carolina University, Kansas State University, Michigan State University, Purdue University, Rutgers University, South Dakota State University, Syracuse University, Tuskegee University, University of Florida, University of New Hampshire, University of Rhode Island, University of Wisconsin-Madison, and West Virginia University. The intervention was grounded in the PRECEDE-PROCEED model of health promotion and CBPR. Full-time undergraduate students aged 18–24 years with reliable internet access were eligible. Exclusion criteria included enrollment in a health-related major or a nutrition course, BMI <18.5 kg/m², pregnancy, or presence of a life-threatening illness. Recruitment was conducted via campus listservs, flyers, and newspaper advertisements. Of 6,277 screened students, 1,639 enrolled at baseline.

Participants were randomized to either the 10-week YEAH intervention or a control group. The intervention included 21 virtual lessons (1–3 per week) delivered via the YEAH website, supported by personalized “nudges” (messages and videos tailored to readiness to change) sent 4 times per week. During the subsequent 12-month follow-up, nudges were reduced to 4 per month. The control group did not receive program content during the intervention.

Primary outcomes included body weight, height, BMI and fruit and vegetable intake. Assessments were conducted at baseline, post-intervention (3 months), and follow-up (15 months). Anthropometrics were measured in person by trained staff, while surveys captured diet, physical activity, stress, sleep, and readiness to change. All anthropometric assessments were recorded twice, and averages were reported. Weight and waist circumference were rounded to the nearest 0.1 kg and 0.1 cm, respectively.

### Get FRUVED Trial

Get FRUVED was a 24-week, peer-led hybrid intervention with data collected in five waves. Data collected from wave 1 conducted in Fall 2015 and Spring 2016 across 8 US universities, with follow-up assessments in Spring 2016 was used for this analysis (24). Eligibility criteria for undergraduate participants included low fruit and vegetable intake (<2 cup equivalents fruit and/or <3 cup equivalents vegetables daily) and at least one risk factor for weight gain in the first year of college (e.g., parental overweight/obesity, first-generation college status, racial/ethnic minority, low-income, or BMI >25 kg/m²). Of 2,757 screened students, 1,149 enrolled at baseline. The study was approved by the Institutional Review Boards of all participating universities, and participants provided written informed consent.

Participants were randomized to either the FRUVED intervention or a control group. Intervention sites included the University of Florida, University of Tennessee, West Virginia University, and South Dakota State University, and control sites included Syracuse University, Kansas State University, University of Maine, and Auburn University. The intervention combined weekly online content (goal setting, event calendars, educational materials) with twice-weekly in-person campus events. Smaller midweek events reinforced weekly health themes, while larger Friday events offered interactive, hands-on learning. Peer leaders (trained upperclassmen) and research partners coordinated digital materials, social media, and campus activities. Control participants did not receive intervention programming.

Primary outcomes included fruit and vegetable intake, weight, BMI, and psychosocial and behavioral measures relevant to college health. Anthropometrics were measured in person by the research team, while surveys captured diet, physical activity, stress, and sleep. Assessments of participants in wave 1 occurred at baseline (Fall 2015), Spring 2016, Fall 2016, Spring 2017, and Spring 2018. For a detailed overview of YEAH and FRUVED lesson topics and outcomes targeted, see **Supplementary Table 1**.

#### Outcomes Assessment

For this secondary analysis, common outcomes from the YEAH and FRUVED datasets at the baseline and post-study time-point (YEAH: month 3; FRUVED: end of second semester) were merged. These included anthropometrics (weight, waist circumference), dietary data, physical activity, sleep, and perceived stress. A detailed list of study variables, including independent and dependent measures, with corresponding assessment procedures and references is provided in **Supplementary Table 2**. Dietary intake variables were assessed using the National Cancer Institute’s Fruit and Vegetable and Fat screeners (25,26). Items related to frequency of consumption were converted to frequency per day using the National Cancer Institute’s scoring algorithm (27,28). Items related to quantity were recoded to the exact numeric value listed (i.e., ½ cup = 0.5), or the average if provided as a range. For quantity items worded as “more than,” or “less than,” values were rounded up or down by 0.5 or 0.25 cups, respectively (i.e., less than ½ cup = 0.25, more than 2 cups = 2.5). To calculate cups/day of fruits and vegetables, frequency per day values were multiplied by recoded quantity values. NCIFV scores were calculated by summing cups/day values of fruit and vegetables. Total fruit and fruit juice were calculated by summing cups/day of whole fruit and fruit juice variables. Comparatively, grams/day of dietary fat variables were calculated by multiplying frequency per day values by sex-specific median serving sizes (grams) using National Cancer Institute Fat Screener scoring procedures (28).

Percentage of energy from fat was calculated using a National Cancer Institute regression equation, which included dietary fat variables and estimated regression coefficients. Total processed meat was quantified as the sum of grams/day values for bacon/sausage and beef/pork hot dogs. Physical activity was measured with the short form International Physical Activity Questionnaire (IPAQ) (29). Sleep was self-reported as average hours per night in YEAH and at baseline in FRUVED, while at the follow-up assessment in FRUVED, sleep duration was calculated from reported bedtimes, sleep onset, and wake times. Perceived stress was measured using the 14-item Cohen’s Stress Scale (30). Items from the perceived stress questionnaire were assessed using a 5-point Likert scale, ranging from never (0) to almost always (4). Positively phrased items were reverse-scored (i.e., 4-0) for quantification of total perceived stress, which was calculated as the sum of all 14 perceived stress items (31). The negatively phrased items were summed to yield the negative stress subscale, while the positively phrased items were summed to generate the positive stress subscale.

#### Statistical Analyses

Statistical analyses were conducted in R (version 4.5.2). Secondary analyses were performed using baseline-adjusted linear regression models using the lm() function in R to evaluate post-intervention outcomes in the combined sample (FRUVED and YEAH). Fixed effects models included experimental group (intervention, control), sex (female, male), and either race (White, Black, Other) or Hispanic ethnicity (yes, no), along with their two- and three-way interaction terms, with Group included in all interaction models (e.g., Group x Sex, Group x Ethnicity,

Group x Sex x Ethnicity for ethnicity model and Group x Sex, Group x Race, Group x Sex x Race for race model). Due to extremely small sample sizes in racial categories other than White and Black, these were combined to form the “Other” category. To account for initial differences between groups as well as differences by the study type and state of data collection, all models were adjusted for the baseline value of the respective outcome, baseline BMI, study, and state.

Missing data were addressed prior to analysis using multiple imputation by chained equations implemented with the mice package using predictive mean matching (32). Imputation models were specified separately for the race-based and ethnicity-based analyses. For race models, the imputation model included Age, BMI, University, Dataset, Race, School Year, and the respective baseline value of each outcome as predictors. For Ethnicity models, Race was replaced with Hispanic ethnicity as a predictor. The number of imputations was determined using the von Hippel rule based on the maximum observed proportion of missing data (33). Imputed datasets were generated with 50 iterations per chain, analyses were conducted within each imputed dataset, and parameter estimates were pooled using Rubin’s rules (34). Estimated marginal means, planned contrasts, and joint tests of model terms were computed using the emmeans package from pooled model estimates (35). P-values for pairwise comparisons were adjusted for multiple testing using the multivariate t (mvt) method (36).

Model assumptions were evaluated using residual diagnostics, including skewness and kurtosis statistics, Levene’s tests, and graphical inspection of residual distributions. Variables not meeting assumptions were transformed using Johnson’s family of transformations in JMP Pro (version 17.0). When the transformed model better satisfied distributional assumptions, but both transformed and non-transformed models yielded the same statistical conclusions, then non-transformed values and results are presented for interpretability. For outcomes reported in “units,” values represent transformed variables rather than raw measurements.

## RESULTS

The demographic characteristics of the participants are shown in **Table 1**. All outcomes reflect post-study values adjusted for their baseline values, baseline participant BMI, study (YEAH vs FRUVED), and state.

**Table 1.** Baseline Participant Demographics Characteristics of Young College Adults.

|  |  | **Group** |  |
| --- | --- | --- | --- |
| Age<sup>a</sup> |  | **Control (N=1330)** | **Intervention (N=1458)** |
|  |  | 18.92 ± 1.12 | 18.82 ± 1.08 |
| Sex<sup>b</sup> | Female | 860 (66.4%) | 976 (68.6%) |
|  | Male | 449 (33.7%) | 462 (31.4%) |
| Race | Black or African American | 160 (12.7%) | 184 (13.5%) |
|  | *Female* | 106 (8.5%) | 136 (10.0%) |
|  | *Male* | 53 (4.3%) | 47 (3.5%) |
|  | White | 879 (69.2%) | 969 (70.6%) |
|  | *Female* | 584 (46.8%) | 666 (49.0%) |
|  | *Male* | 280 (22.4%) | 294 (21.6%) |
|  | Other | 227 (18.0%) | 217 (16.0%) |
|  | *Female* | 138 (11.1%) | 131 (9.6%) |
|  | *Male* | 87 (7.0%) | 86 (6.3%) |
| Ethnicity | Hispanic | 126 (9.6%) | 172 (12.1%) |
|  | *Female* | 75 (5.8%) | 126 (8.9%) |
|  | *Male* | 49 (3.8%) | 46 (3.2%) |
|  | Non-Hispanic | 1190 (90.5%) | 1261 (88.0%) |
|  | *Female* | 778 (60.0%) | 840 (59.0%) |
|  | *Male* | 396 (30.5%) | 411 (28.9%) |

<sup>a</sup>Mean ± SD, <sup>b</sup>(N %)

### Anthropometrics

Body weight and waist circumference demonstrated significant Group x Sex x Ethnicity interactions (**Table 2**). Among Hispanic males, intervention participants had higher body weight than control participants (+3.8 kg) (p < 0.05). Hispanic males in the intervention group also had higher body weight and waist circumference than Hispanic females (+3.7 kg, +0.40 units) and non-Hispanic males (+3.7 kg, +0.32 units) (p < 0.05).

**Table 2.**
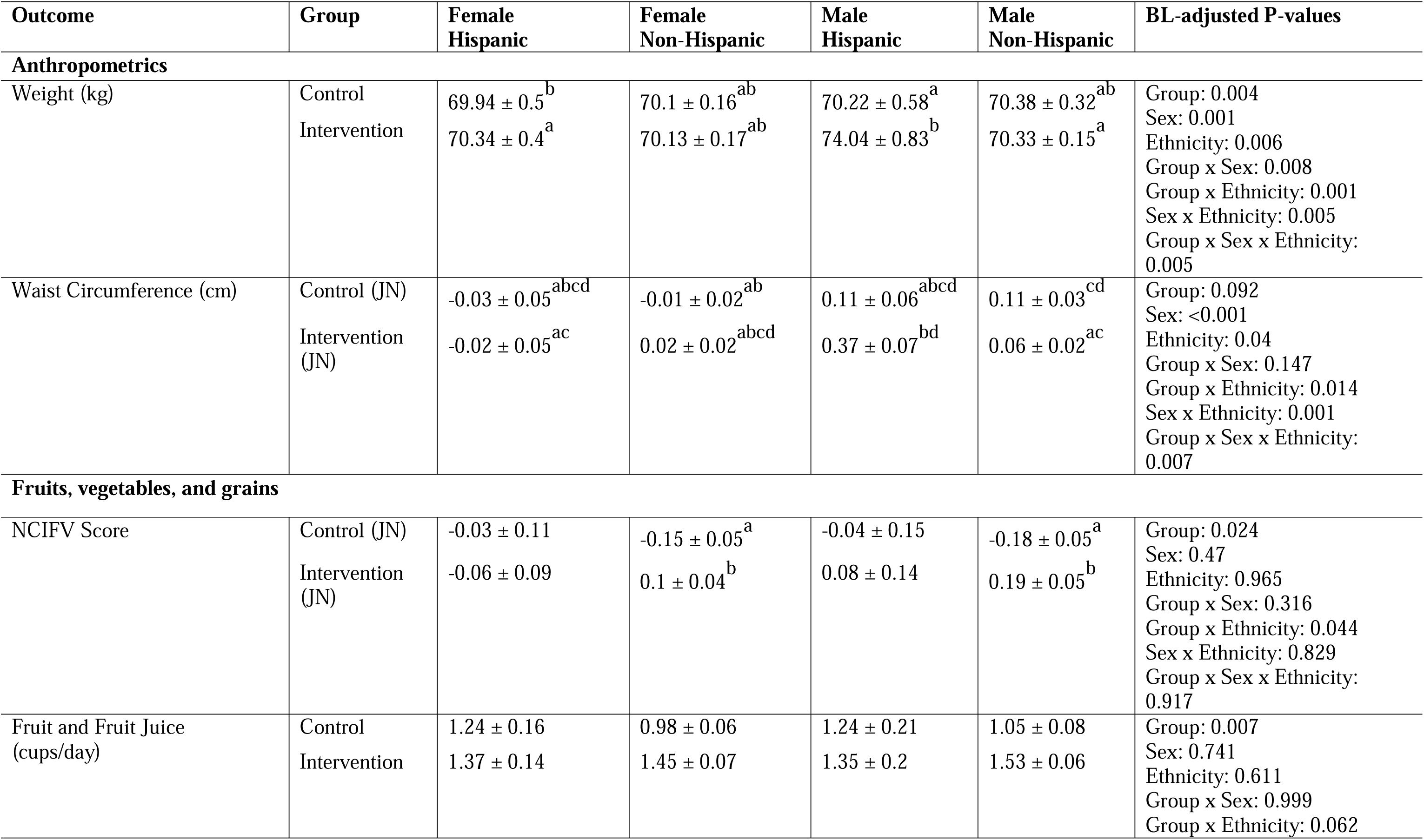
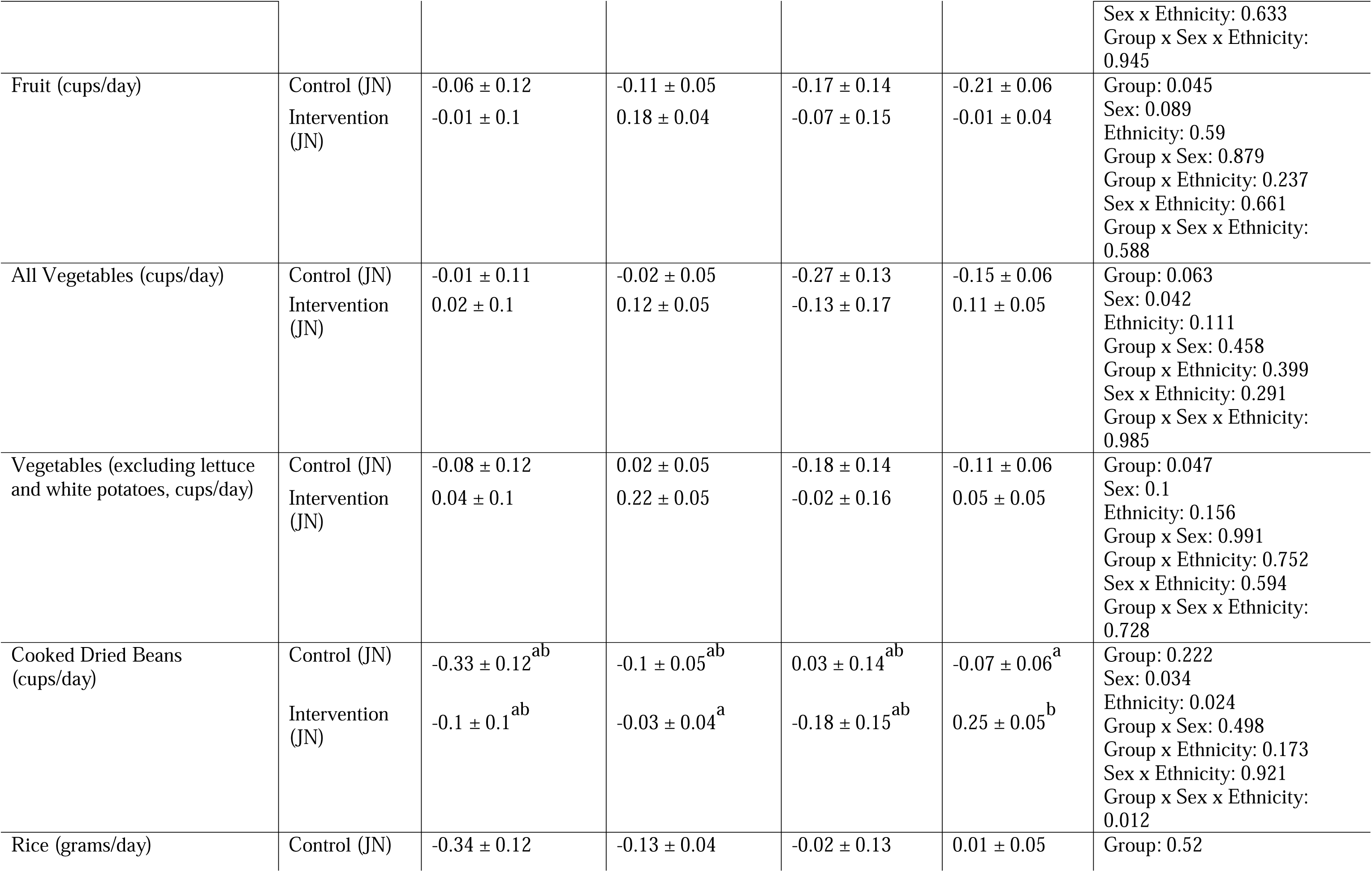
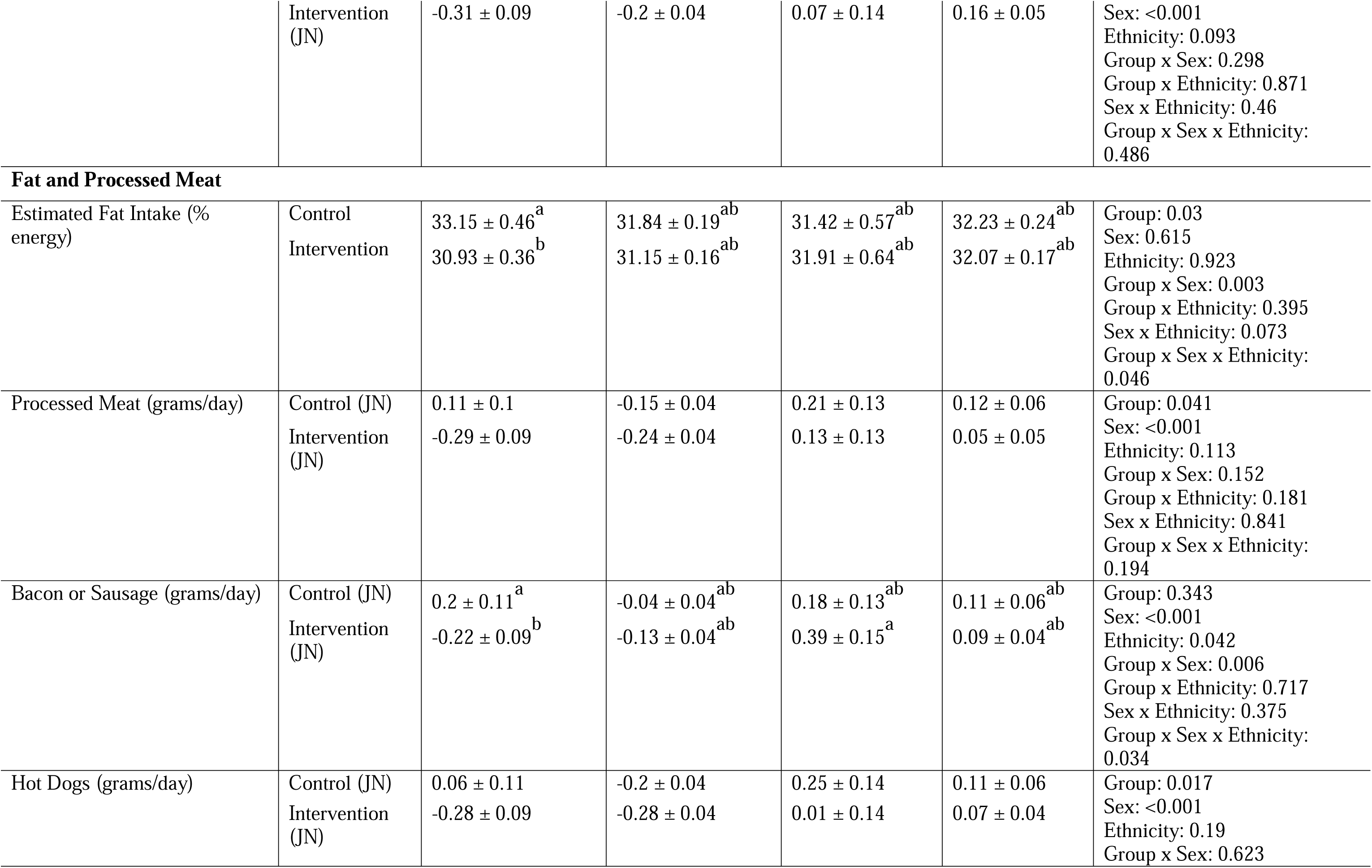
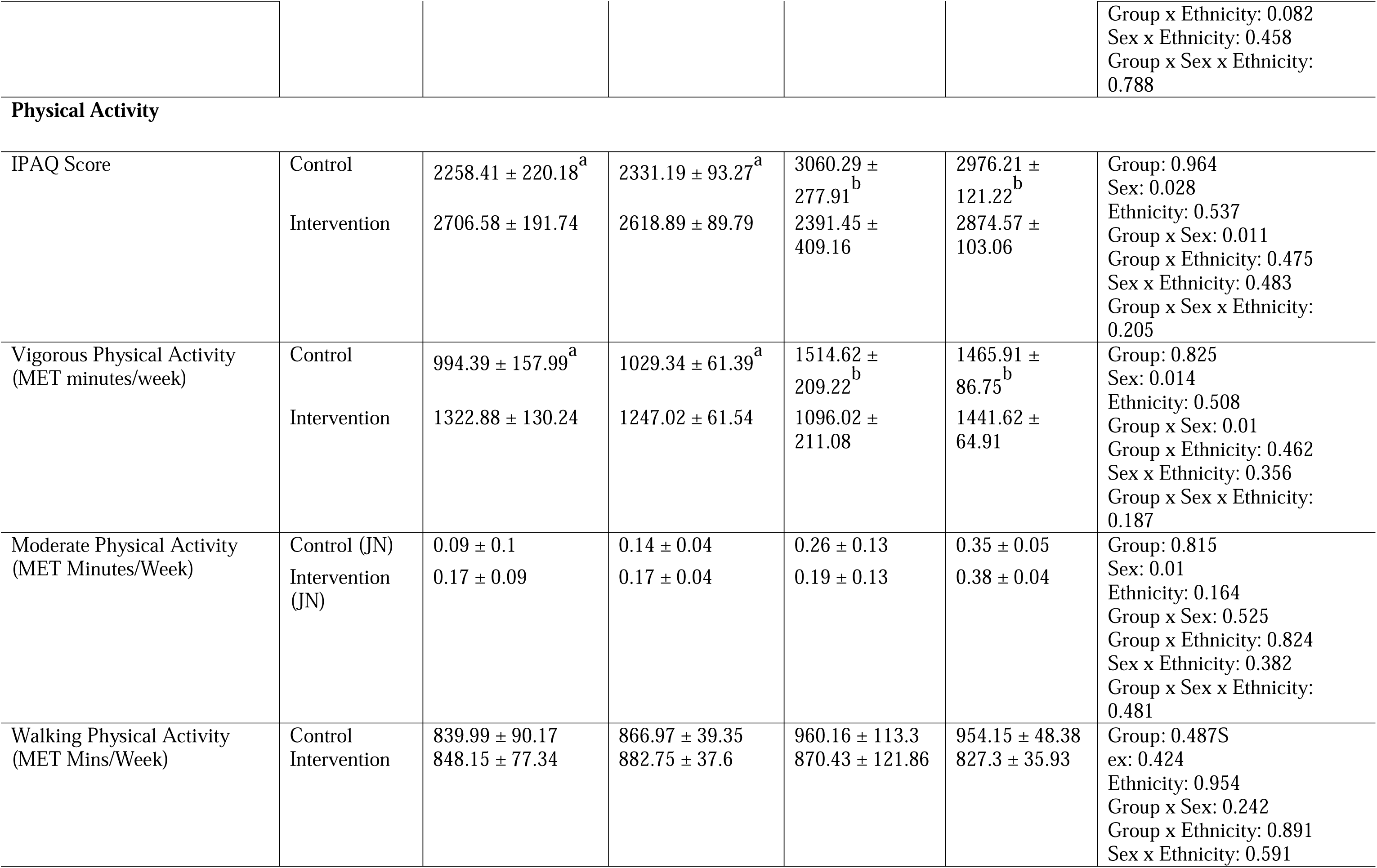
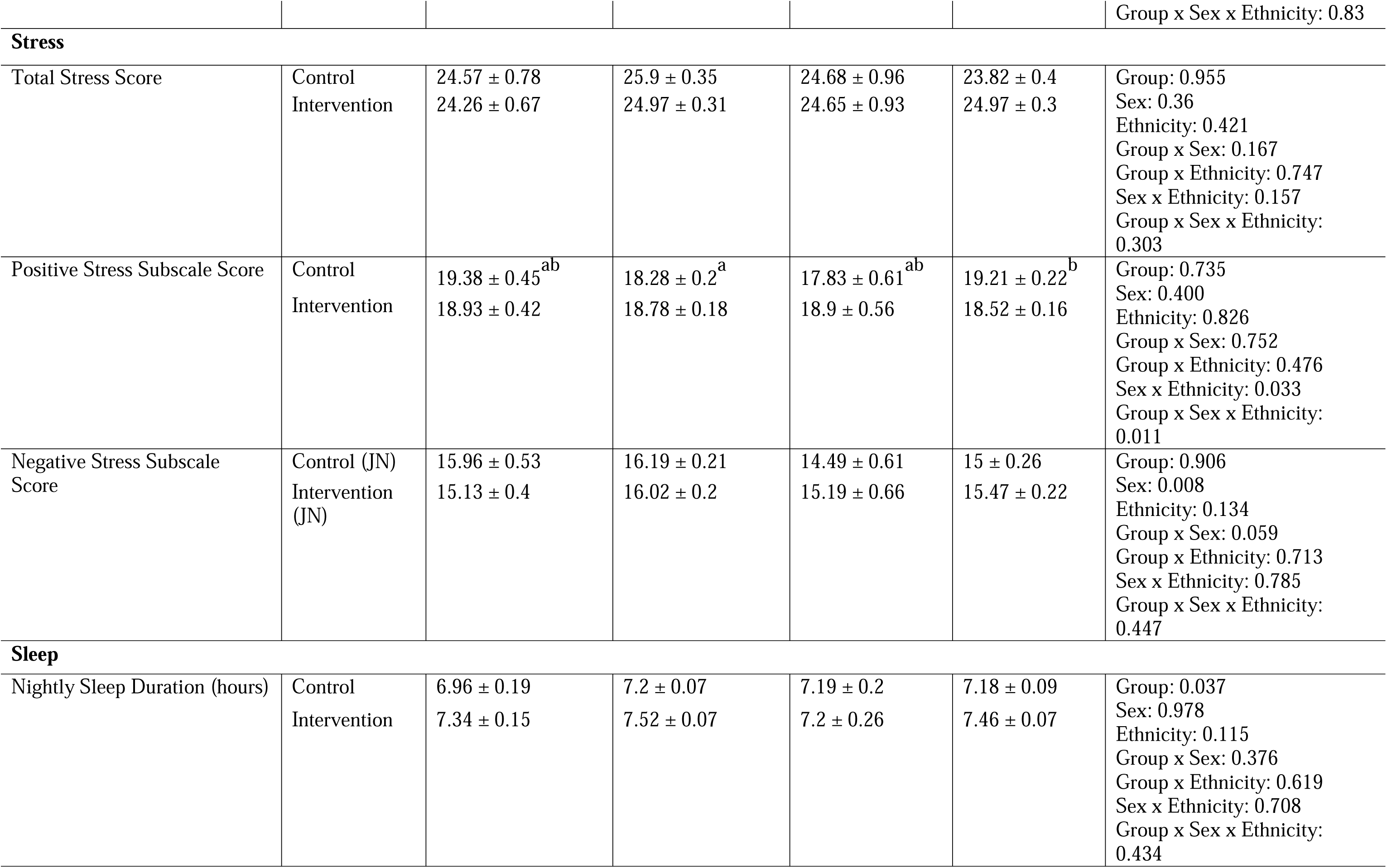
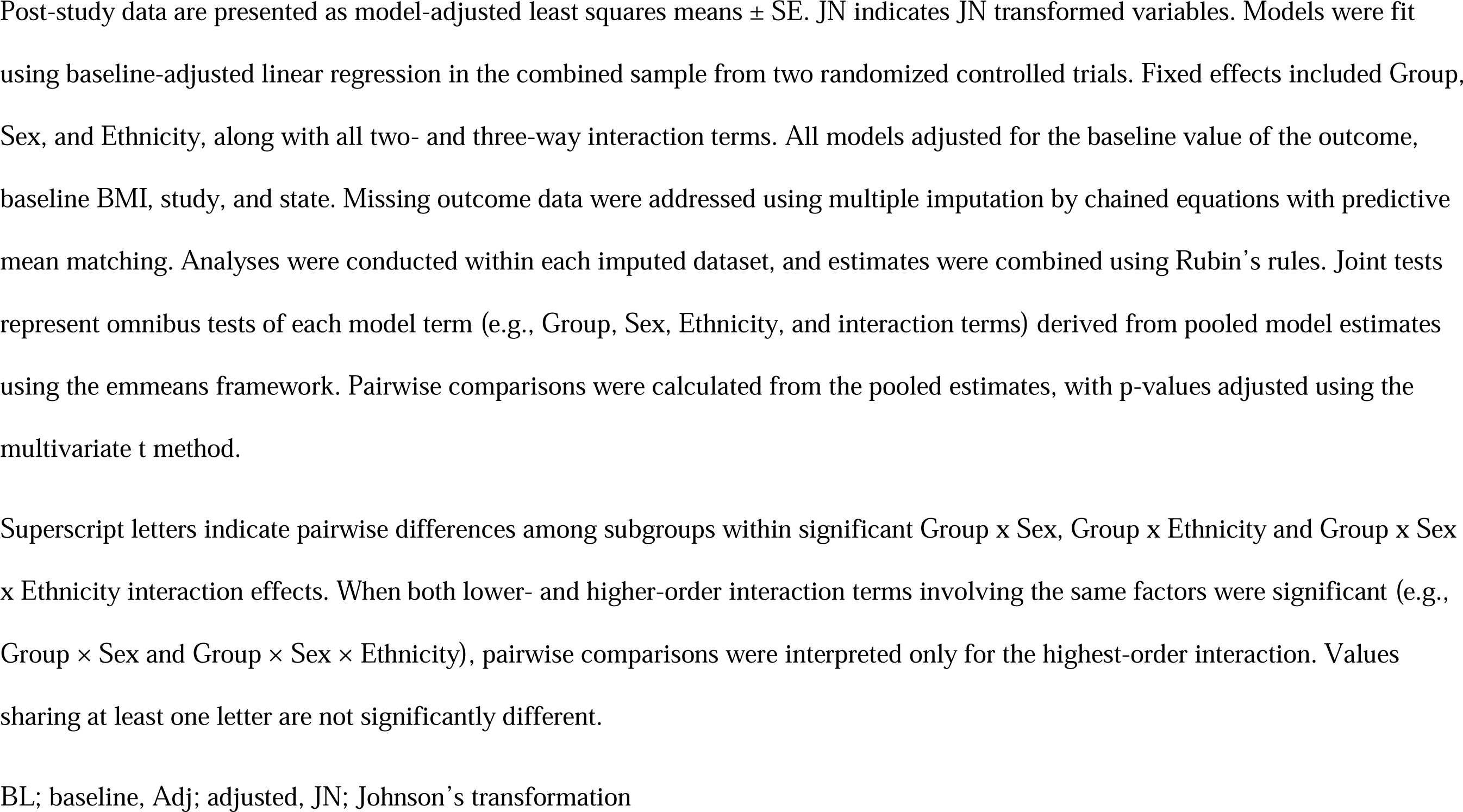
Model-Adjusted Group, Sex, and Ethnicity Effects on Post-Study Outcomes in Young College Adults.

### Dietary Intake

#### Fruits, vegetables, and grain intake

A significant Group x Ethnicity interaction was observed for the NCI fruit and vegetable (NCIFV) score (**Table 2**). Among non-Hispanic participants, intervention participants had higher NCIFV scores than control participants (+0.31 units) (p < 0.05). In the race model, no significant Group x Race interaction was observed for NCI fruit and vegetable scores (**Table 3**). However, intervention participants had higher overall NCI fruit and vegetable scores (+0.27 units) than control participants (Group main effect, p < 0.05) (**Table 3**).

**Table 3.**
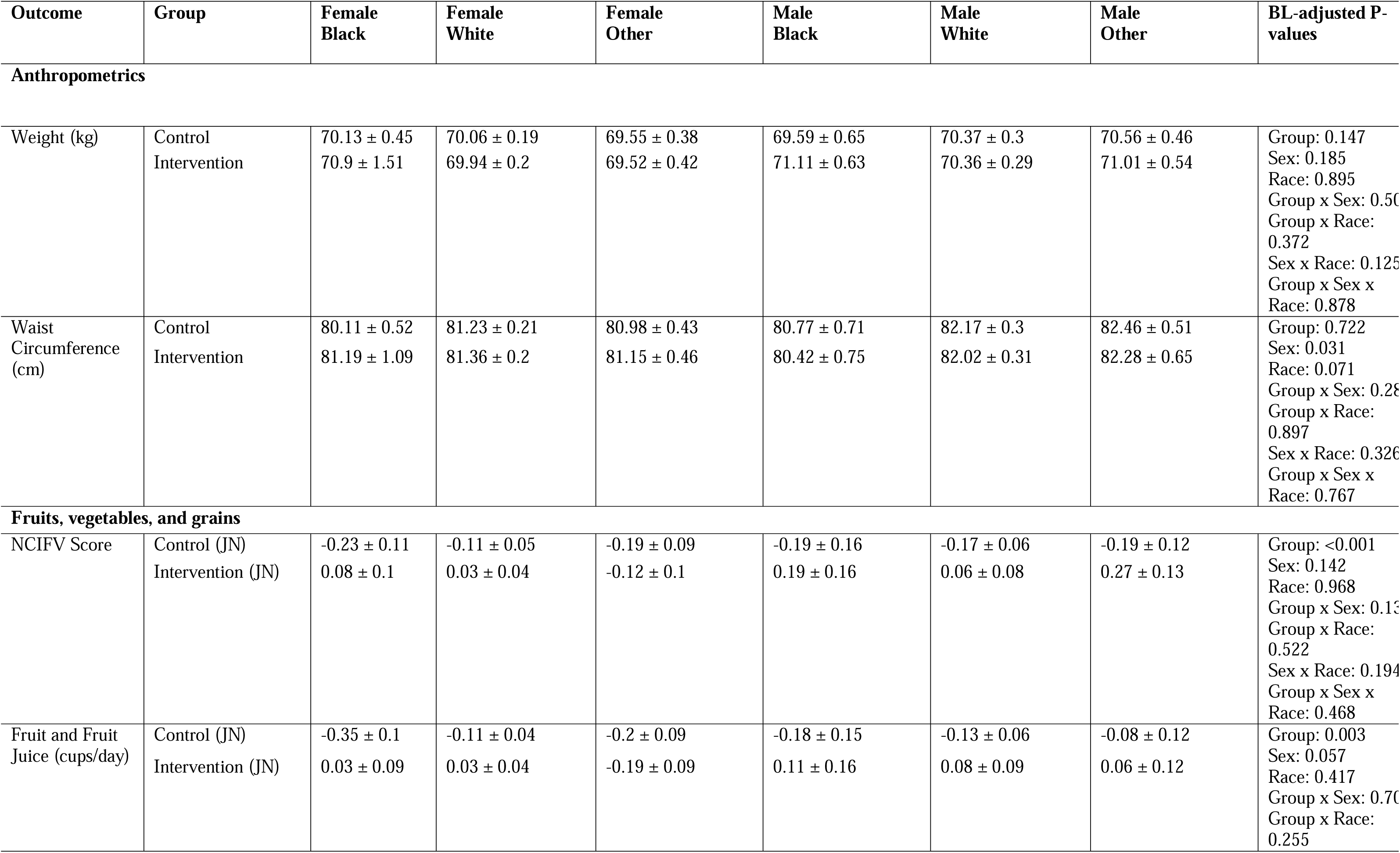
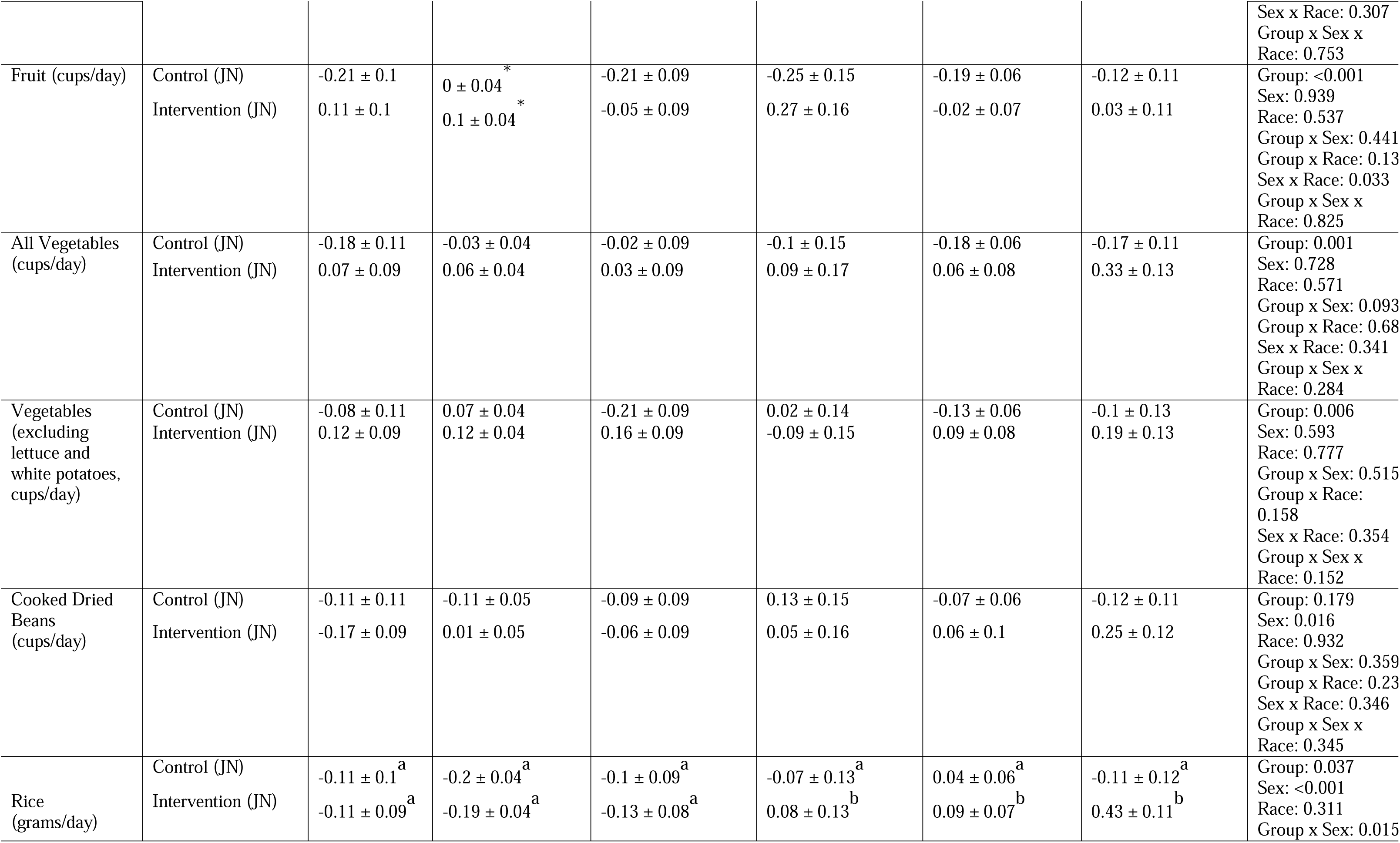
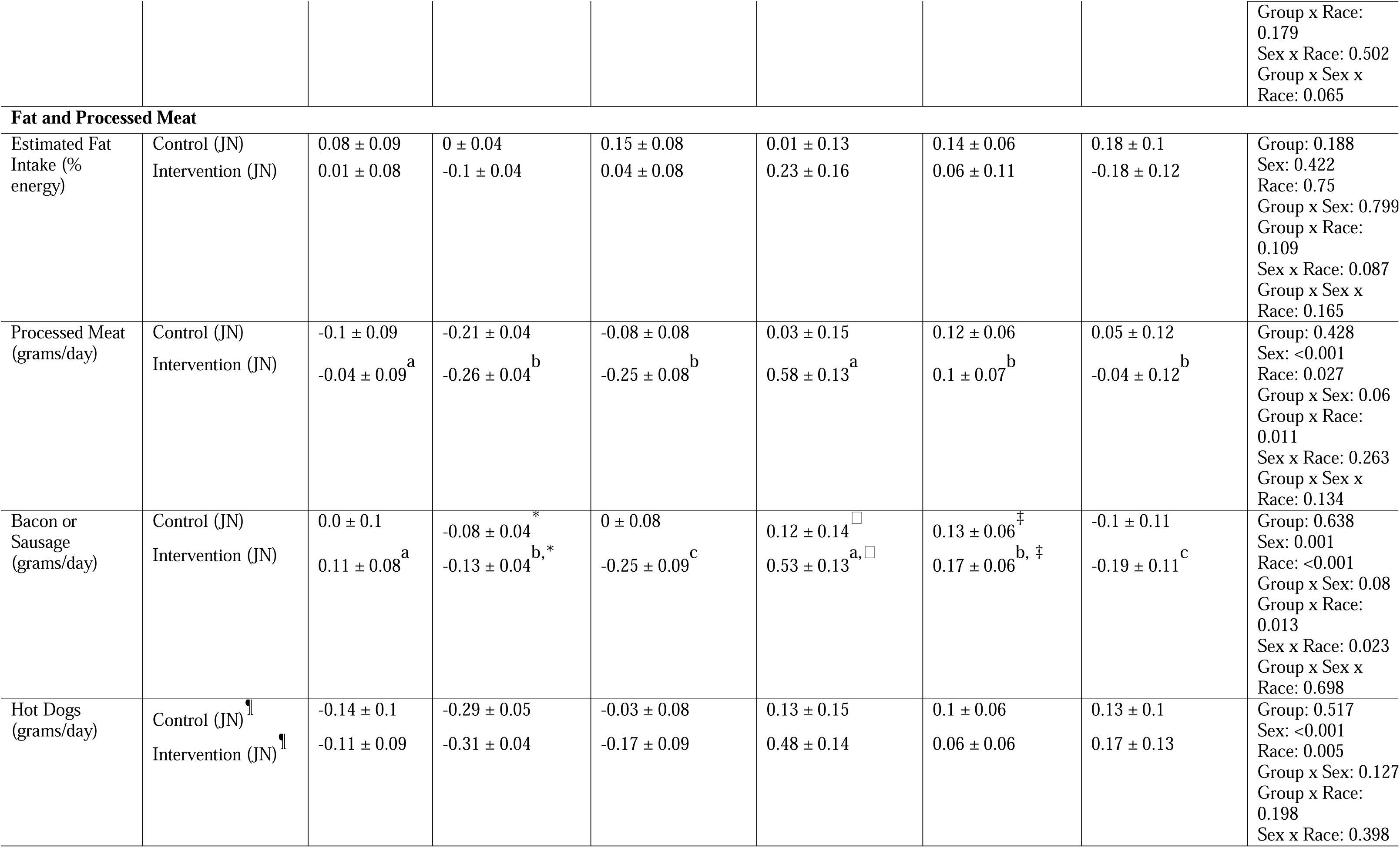
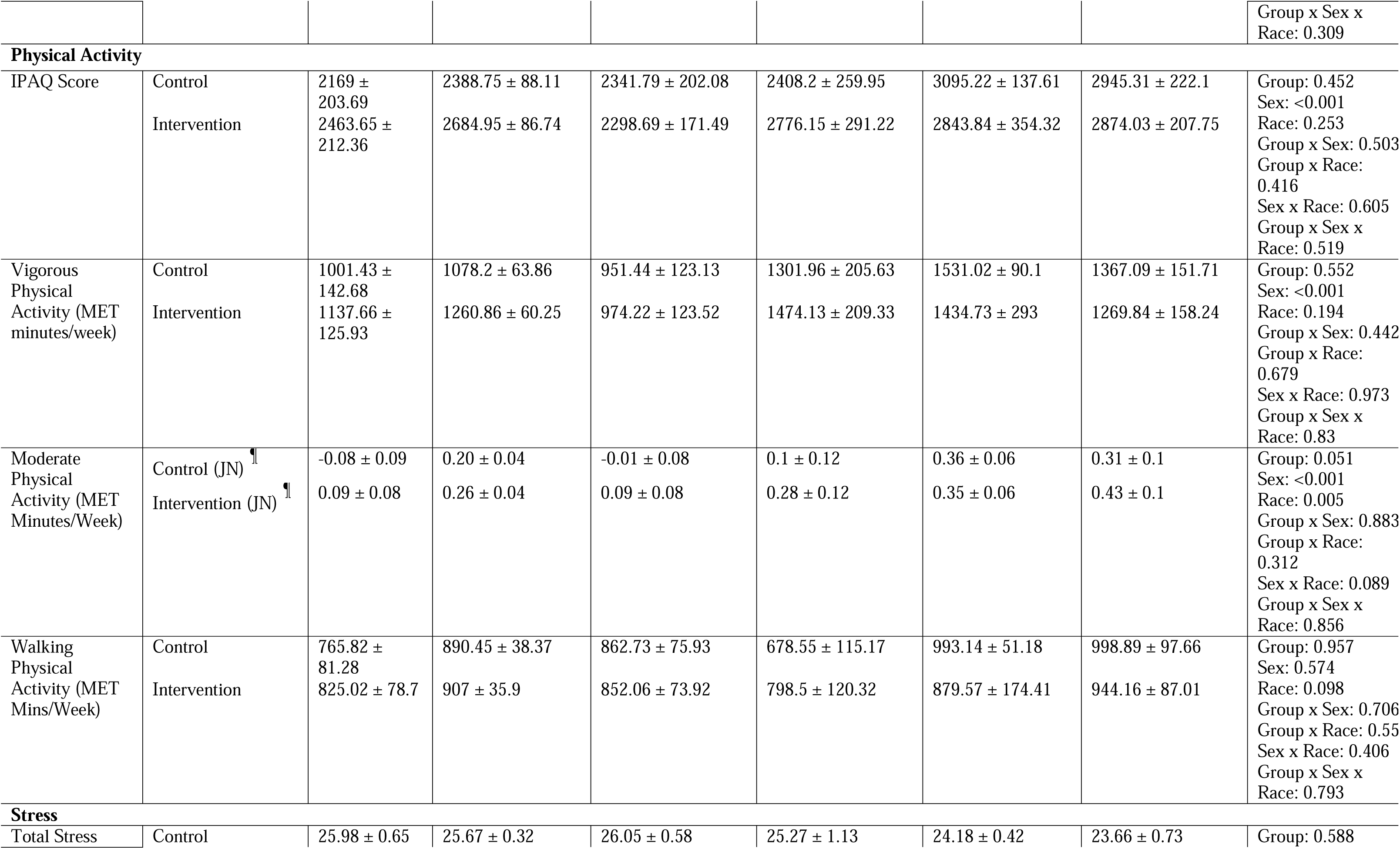
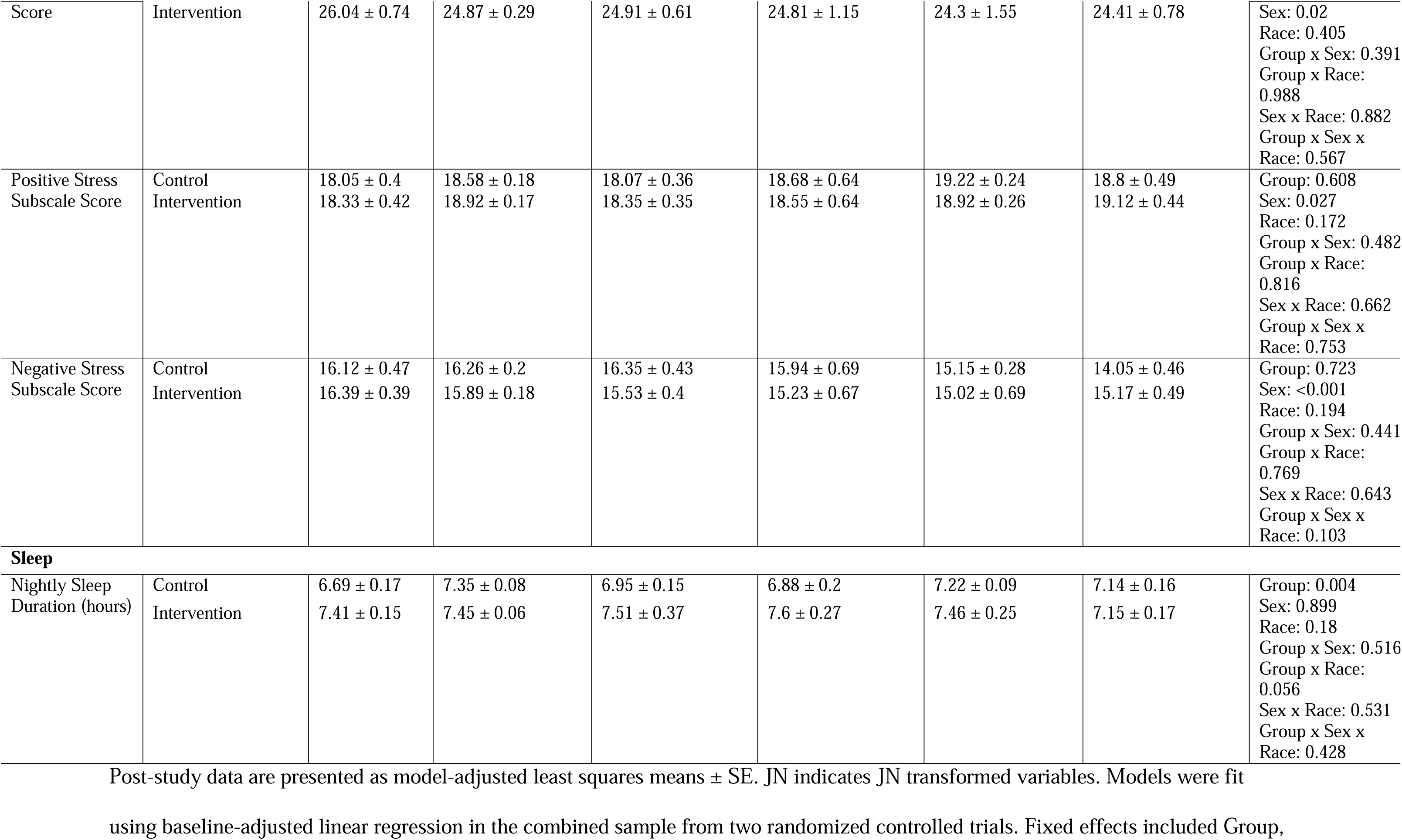
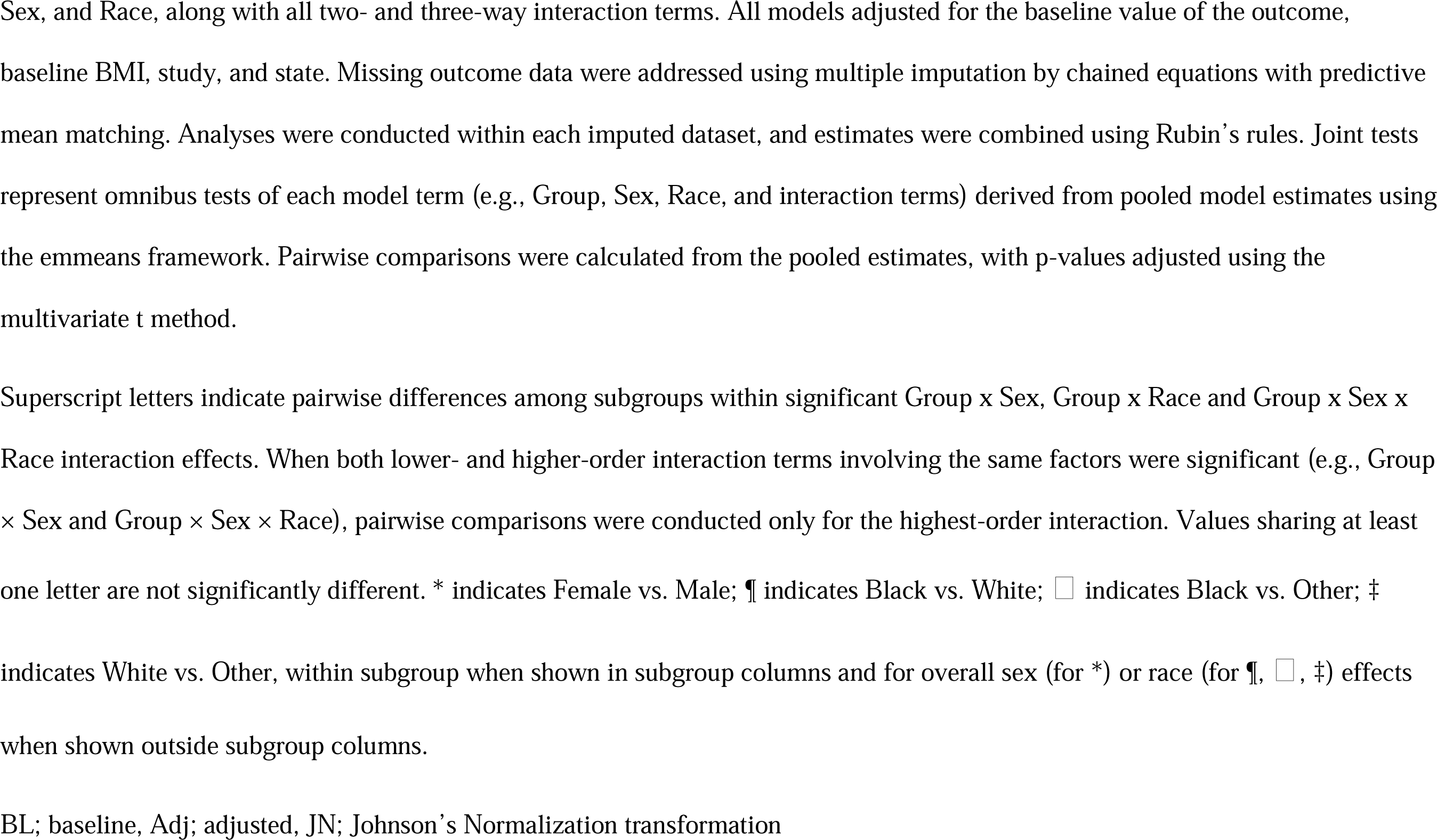
Model-Adjusted Group, Sex, and Race Effects on Post-Study Outcomes in Young College Adults.

Significant Group x Sex x Hispanic interactions were observed for cooked dried beans (**Table 2**) where non-Hispanic intervention males consumed more beans than their control counterparts (+0.32 units; p < 0.05) and their non-Hispanic female intervention counterparts (+0.28 units; p < 0.05).

In both the ethnicity and race models, several dietary outcomes demonstrated significant Group main effects regardless of ethnicity or race. Intervention participants consumed more total fruit (+0.16 units, ethnicity model; +0.24 units, race model), and more other vegetables (+0.16 units, ethnicity model; +0.17 units, race model) than control participants (p < 0.05) (**Tables 2-3, Supplementary Tables 5-6**).

In the race model, rice intake demonstrated a significant Group x Sex interaction (**Table 3**). Within the intervention group, females consumed less rice than males (−0.34 units) (p < 0.05). Among males, intervention participants consumed more rice than control participants (+0.25 units) (p < 0.05).

#### Processed meat and fat intake

Significant Group x Sex x Hispanic interactions were observed for bacon/sausage intake and percent energy from fat (**Table 2**). Compared to their male counterparts, Hispanic females consumed less bacon/sausage (−0.61 units) in response to the intervention. Furthermore, Hispanic females in the intervention group ate less bacon/sausage (−0.41 units) and less energy from fat (−2.23%) compared to their control counterparts (p < 0.05) as well. However, regardless of sex and ethnicity, intervention participants consumed fewer hot dogs (−0.17 units) and processed meat (−0.16 units) than control participants (Group main effect, p < 0.05) (**Tables 2-3**).

Significant Group x Race interactions were observed for bacon/sausage and processed meat intake (**Table 3**). Within the intervention group, Black participants consumed more bacon/sausage and total processed meat than White participants (+0.30 units; +0.35 units) and participants classified as Other race (+0.54 units; +0.42 units) (p < 0.05).

#### Physical Activity

In the ethnicity model, physical activity outcomes demonstrated significant Group x Sex interaction effects (**Tables 2-3**). Within the control group, females reported lower IPAQ score (−723 MET-min/week) and vigorous physical activity (−0.478 MET-min/week) than males (p < 0.05). In the race model, while no interaction effects of race and sex with group were found, there were overall significant sex and race effects for selected physical activity outcomes (**Table 3, Supplementary Tables 3-6**). Specifically, males reported significantly higher IPAQ scores (+329 MET-min/week) (p <0.05) than females and Black participants reported lower moderate physical activity (−0.19 units) than their White peers (p < 0.05).

#### Sleep

In both ethnicity and race models, nightly sleep duration demonstrated a significant Group main effect (**Tables 2**-3, **Supplementary Tables 3-6**). Intervention participants reported 0.25 hours (ethnicity model) and 0.39 hours (race model) more sleep per night than control participants (p < 0.05).

#### Stress

In the ethnicity model, negative stress scores (perceived distress) demonstrated a significant main effect of Sex, while positive stress scores (coping ability) demonstrated a significant Group x Sex x Ethnicity interaction (**Table 2, Supplementary Tables 3 and 5**). Females reported higher negative stress (perceived distress) than males (+0.79 units, p < 0.05). Within the control group among non-Hispanic participants, females reported lower positive stress (coping ability) than males (−0.92 units, p < 0.05). In the race model, stress outcomes demonstrated only a significant effect by sex (**Table 3, Supplementary Tables 4 and 6**). Females reported higher negative stress (perceived distress; +0.99 units), higher total stress (+1.15 units), and lower positive stress (coping ability; −0.50 units) than males (all p < 0.05).

**Supplementary Tables 3 and 4** show all outcomes for the ethnicity and race models respectively, including the non-transformed data for interpretation of values when the primary analysis was performed on transformed variables. **Supplementary Tables 5 and 6** show the model-adjusted means ± SE for the overall group, sex, and ethnicity/race effects from the respective ethnicity and race models for variables.

## DISCUSSION

The YEAH and GetFruved studies were CBPR research trials conducted among college undergraduates across multiple university campuses. Both interventions used non-diet approaches focused on obesity and weight gain prevention, delivered through peer-led social marketing strategies (GetFruved) (37) or web-based training initiatives (YEAH) (38). The present secondary analysis examined whether the effectiveness of these interventions was moderated by the intersecting influences of sex, ethnicity, and race, factors that evidence has consistently shown to be associated with lifestyle behaviors (1,18–20,38). Our findings reveal substantial heterogeneity in intervention-related changes in dietary behaviors across these demographic factors. In contrast, sex, race, or ethnicity did not moderate the effect of the intervention on physical activity, sleep, or perceived stress outcomes. With respect to anthropometric outcomes, the intervention appeared to elicit minimal responsiveness for body weight maintenance among Hispanic males.

Non-Hispanic participants in the intervention group improved intake of fruits and vegetables. This may be especially relevant given national data showing declines in overall diet quality among non-Hispanic White adults, suggesting that interventions such as these could help offset unfavorable dietary trends in this population (20). With regards to Hispanic populations, prior research has shown that fruit and vegetable consumption patterns vary considerably depending on cultural background, acculturation status, and socioeconomic and food environment context (39–41), and in some cases Hispanic individuals may have had higher legume and/or vegetable intake compared with non-Hispanic peers (18,42). If baseline intake was comparatively higher in certain components, ceiling effects could have limited the possibility of detecting change. Alternatively, intervention messaging and food examples may have aligned more closely with dietary practices common among non-Hispanic participants, thereby facilitating greater behavioral adoption in that group.

Sex by ethnicity differences were also found in processed meat and fat related outcomes within the intervention group. NHANES 2015–2016 documents higher processed meat intake among men than women (43). In the present study, specifically, within Hispanic participants, reductions in bacon/sausage intake and percent energy from fat were evident among females but not males, suggesting that processed meat reduction may be less responsive among Hispanic young adult men under broadly delivered lifestyle programming and may benefit from sex and culturally relevant tailoring. This supports a focus on young adult men as a priority group for processed meat reduction and suggests that interventions may need added, subgroup-relevant supports to produce comparable change among Hispanic males.

Overall, dietary changes were less favorable among Hispanic participants, and particularly among Hispanic males, with limited improvement in fruit and vegetable intake among Hispanic participants and smaller reductions in processed meat intake among Hispanic males. Consistent with this pattern, Hispanic males also exhibited higher body weight and adiposity than non-Hispanic males and higher body weight than Hispanic females, suggesting reduced responsiveness to the intervention related to weight in this subgroup. Together, these findings indicate that fruit and vegetable promotion and processed meat reduction strategies may not be uniformly effective across ethnic and sex subgroups. Interventions targeting young adults may therefore benefit from culturally and sex-relevant tailoring to improve diet quality and downstream health outcomes.

Processed meat intake also showed race-specific patterning in our intervention sample, with Black participants reporting higher total processed meat intake than White and Other race participants. This pattern suggests that broadly delivered intervention content may have been less effective at shifting processed-meat behaviors for some groups, or that contextual factors (e.g., dining hall offerings, meal plans, cost constraints, time demands, and nearby food options) continued to shape intake despite exposure. Nationally representative NHANES data provide useful context but indicate a different adult pattern: in 2015–2016, non-Hispanic White adults reported the highest processed meat intake overall (43). Findings from the Diabetes Prevention Program Outcomes Study (DPPOS) are consistent with this adult pattern, with higher red meat intake among non-Hispanic White participants than African American, Hispanic, and Asian American participants in the lifestyle arm (44). NHANES is nationally representative of U.S. adults, and DPPOS enrolled adults with overweight/obesity and elevated diabetes risk participating in a structured lifestyle trial. In contrast, our sample consists of college students whose food choices are shaped by campus dining systems, meal plans, and the surrounding campus food environment. The persistence of higher processed meat intake among Black participants is notable given evidence that genotypic susceptibility to processed meat–related colorectal cancer risk may be higher among this population (45). Although we did not assess genotype or clinical outcomes, this literature highlights the importance of achieving equitable reductions in processed meat intake during young adulthood.

Behavioral outcomes beyond dietary intake showed a mixed pattern of responsiveness. Sleep duration was higher among intervention participants regardless of race and ethnicity categorization. This aligns with existing evidence that multicomponent interventions often benefit sleep quality likely due to the synergistic effects of diet, exercise, and other behavioral components (46,47). In contrast, physical activity did not demonstrate a measurable intervention effect, suggesting that meaningful changes in physical activity in college-aged populations may require more structured, dose-specific programming. Perceived stress patterns reflected sex differences rather than intervention responsiveness, with females reporting higher stress overall, aligning with literature documenting sex-differentiated stress and responsivity during emerging adulthood (48,49). As with physical activity, addressing psychosocial stressors, especially among college-aged females, may benefit from targeted stress-management or mental wellness components to strengthen the overall health impact of future interventions.

A notable strength of this study was the large sample size across multiple states in the US, which enhanced statistical power to detect differences across racial and ethnic subgroups. In addition, we used conservative multiple post-hoc testing adjustments for sex, race, and ethnicity comparisons, strengthening the robustness and credibility of the subgroup findings. However, a limitation of the present analysis was the inability to examine smaller racial categories classified as “Other” or categories within Hispanic and non-Hispanic categories due to limited sample size. An additional limitation of the present study was the small sample size of Hispanic males relative to other groups. Another limitation was the difference in intervention duration between YEAH (3 months) and FRUVED (9 months), which may have introduced variability in intervention exposure. However, to account for potential study-based differences, study was included as a covariate in all models. Moreover, it is possible that some dietary intake changes were not captured because a comprehensive food frequency questionnaire or repeated dietary recalls were not administered. Lastly, we were unable to assess the intersection of food security status with race or ethnicity in this combined analysis because food security data was not available in the YEAH study.

Collectively, the findings from this analysis highlight two central implications. First, multi-component lifestyle interventions can produce measurable improvements in fruit and vegetable intake and reductions in processed meat intake among young adults, although these intervention effects are not uniform across demographic groups. Second, the persistence of race and ethnicity subgroup differences despite adjustment for baseline characteristics and study context underscores the importance of intersectional analyses in behavioral nutrition research. Future work should focus on refining intervention components to enhance responsiveness among subgroups in whom dietary change does not fully translate into health improvements.

## Data Availability

All data produced in the present study are available upon reasonable request to the authors

## Acknowledgment

The YEAH project was supported by National Research Initiative Grant (2009-55215-05460) and the GetFruved project was supported by Agriculture and Food Research Initiative Grant (2014-67001-21851) from the USDA National Institute of Food and Agriculture, *“Get Fruved:” A peer-led, train-the-trainer social marketing intervention to increase fruit and vegetable intake and prevent childhood obesity* –*A2101*. This work was also conducted in collaboration with the USDA Multistate Research Group NC1193, the New Jersey Agricultural Experiment Station, and the New Hampshire Agriculture Experiment Station. The computations for this work were carried out using the high-performance computing infrastructure operated by Research Support Solutions in the Division of Information Technology at the University of Missouri-Columbia; https://doi.org/10.32469/10355/97710.

Post-study data are presented as model-adjusted least squares means ± SE. JN indicates JN transformed variables. Models were fit using baseline-adjusted linear regression in the combined sample from two randomized controlled trials. Fixed effects included Group, Sex, and Ethnicity, along with all two- and three-way interaction terms. All models adjusted for the baseline value of the outcome, baseline BMI, study, and state. Missing outcome data were addressed using multiple imputation by chained equations with predictive mean matching. Analyses were conducted within each imputed dataset, and estimates were combined using Rubin’s rules. Joint tests represent omnibus tests of each model term (e.g., Group, Sex, Ethnicity, and interaction terms) derived from pooled model estimates using the emmeans framework. Pairwise comparisons were calculated from the pooled estimates, with p-values adjusted using the multivariate t method.

## Author Contributions

Conceptualization: all authors; Data analysis: CW, LG, JD; Writing—original draft: CW, LG, JD; Writing—review and editing, all authors.

## Data Accessibility Statement

Data are available upon reasonable request made to the authors. The data are not publicly available due to ongoing analyses.

