## Supplemental Tables for "Demographic Factors Moderate the Effectiveness of Obesity Prevention Interventions: A Secondary Analysis of College Intervention Trials"

**Supplementary Table 1.** YEAH and FRUVED Program Overview and Lesson Topics

| **Intervention** | **Lesson Topics** | **Primary Outcomes Targeted** |
| --- | --- | --- |
| *FRUVED* | - MyPlate - Cardio - Music - Time Management - Meditation - Dance - Fiber - Flexibility - Portions - Strength - Sleep - Positivity - Friendships - Substance Abuse - Body Image - Hydration - Savor Flavor - Try Something New - Maintaining Healthy Lifestyles - Steps | - Weight - Fruit and vegetable intake |
| *YEAH* | - Eating healthy - Eating regularly - Meal planning - Eating a variety - Eating enjoyment - Hunger and fullness - In tune eating - Moving matters - Moving your way - Staying active - Fitness finesse - Mindless eating - Mind and body - Time management - Sleep - Balancing act - Smoking - Body size acceptance - Weight management | - Weight - BMI - Fruit and vegetable intake |

**Supplementary Table 2.** Variables common to the YEAH and FRUVED datasets with their respective assessment methods.

| **Outcome** | **Mode of Assessment** |
| --- | --- |
| BMI | Calculated based on Weight(kg)/Height(m^2^) |
| Height | FRUVED: Measured in-person by trained research staff |
| Weight | FRUVED: Measured in-person by trained research staff |
| Waist Circumference | FRUVED: Measured in-person by trained research staff |
| Dietary Fat Intake | National Cancer Institute Dietary Fat Screener |
| Fruit and Vegetable Intake | National Cancer Institute Fruit and Vegetable Screener |
| Physical Activity | International Physical Activity Questionnaire |
| Stress Perceptions | Cohen’s 14-item Stress Questionnaire |
| Sleep | Study Specific Questionnaire |
| Sex | Study Specific Screener Questionnaire |
| Race | Study Specific Demographic Questionnaire |
| Age | Study Specific Student Life Questionnaire |
| Ethnicity | Study Specific Student Life Questionnaire |
| State | Study Specific Student Life Questionnaire |
| School Year | Study Specific Student Life Questionnaire |

**Supplementary Table 3. Model-Adjusted Group, Sex, and Ethnicity Effects on Post-Study Outcomes in Young College Adults**

| **Outcome** | **Group** | **Female Hispanic** | **Female**  **Non-Hispanic** | **Male Hispanic** | **Male**  **Non-Hispanic** | **BL-adjusted P-values** |
| --- | --- | --- | --- | --- | --- | --- |
| Weight (kg) | Control | 69.94 ± 0.51^ab^ | 70.1 ± 0.16^ab^ | 70.22 ± 0.58^a^ | 70.38 ± 0.32^ab^ | Group: 0.004 Sex: 0.001 Ethnicity: 0.006 Group x Sex: 0.008 Group x Ethnicity: 0.001 Sex x Ethnicity: 0.005 Group x Sex x Ethnicity: 0.005 |
|  | Intervention | 70.34 ± 0.4^a^ | 70.13 ± 0.17^ab^ | 74.04 ± 0.83^b^ | 70.33 ± 0.15^a^ |  |
| Waist Circumference (cm) | Control (JN) | -0.03 ± 0.05^abcd^ | -0.01 ± 0.02^ab^ | 0.11 ± 0.06^abcd^ | 0.11 ± 0.03^cd^ | Group: 0.092 Sex: <0.001 Ethnicity: 0.04 Group x Sex: 0.147 Group x Ethnicity: 0.014 Sex x Ethnicity: 0.001 Group x Sex x Ethnicity: 0.007 |
|  | Intervention (JN) | -0.02 ± 0.05^ac^ | 0.02 ± 0.02^abcd^ | 0.37 ± 0.07^bd^ | 0.06 ± 0.02^ac^ |  |
|  | Control | 80.81 ± 0.56 | 81.19 ± 0.2 | 82.04 ± 0.69 | 82.19 ± 0.32 |  |
|  | Intervention | 81.19 ± 0.48 | 81.4 ± 0.22 | 86.2 ± 0.93 | 81.52 ± 0.2 |  |
| NCIFV Score | Control (JN) | -0.03 ± 0.11 | -0.15 ± 0.05^a^ | -0.04 ± 0.15 | -0.18 ± 0.05^a^ | Group: 0.024 Sex: 0.47 Ethnicity: 0.965 Group x Sex: 0.316 Group x Ethnicity: 0.044 Sex x Ethnicity: 0.829 Group x Sex x Ethnicity: 0.917 |
|  | Intervention (JN) | -0.06 ± 0.09 | 0.1 ± 0.04^b^ | 0.08 ± 0.14 | 0.19 ± 0.05^b^ |  |
|  | Control | 3.15 ± 0.33 | 2.83 ± 0.15 | 2.82 ± 0.4 | 2.71 ± 0.16 |  |
|  | Intervention | 3.2 ± 0.27 | 3.57 ± 0.18 | 3.34 ± 0.42 | 3.77 ± 0.16 |  |
| Fruit and Fruit Juice (cups/day) | Control | 1.24 ± 0.16 | 0.98 ± 0.06 | 1.24 ± 0.21 | 1.05 ± 0.08 | Group: 0.007 Sex: 0.741 Ethnicity: 0.611 Group x Sex: 0.999 Group x Ethnicity: 0.062 Sex x Ethnicity: 0.633 Group x Sex x Ethnicity: 0.945 |
|  | Intervention | 1.37 ± 0.14 | 1.45 ± 0.07 | 1.35 ± 0.2 | 1.53 ± 0.06 |  |
| Fruit (cups/day) | Control (JN) | -0.06 ± 0.12 | -0.11 ± 0.05 | -0.17 ± 0.14 | -0.21 ± 0.06 | Group: 0.045 Sex: 0.089 Ethnicity: 0.59 Group x Sex: 0.879 Group x Ethnicity: 0.237 Sex x Ethnicity: 0.661 Group x Sex x Ethnicity: 0.588 |
|  | Intervention (JN) | -0.01 ± 0.1 | 0.18 ± 0.04 | -0.07 ± 0.15 | -0.01 ± 0.04 |  |
|  | Control | 0.73 ± 0.13 | 0.58 ± 0.05 | 0.48 ± 0.14 | 0.47 ± 0.06 |  |
|  | Intervention | 0.81 ± 0.1 | 0.91 ± 0.07 | 0.77 ± 0.15 | 0.68 ± 0.05 |  |
| All Vegetables (cups/day) | Control (JN) | -0.01 ± 0.11 | -0.02 ± 0.05 | -0.27 ± 0.13 | -0.15 ± 0.06 | Group: 0.063 Sex: 0.042 Ethnicity: 0.111 Group x Sex: 0.458 Group x Ethnicity: 0.399 Sex x Ethnicity: 0.291 Group x Sex x Ethnicity: 0.985 |
|  | Intervention (JN) | 0.02 ± 0.1 | 0.12 ± 0.05 | -0.13 ± 0.17 | 0.11 ± 0.05 |  |
|  | Control | 1.26 ± 0.17 | 1.16 ± 0.08 | 0.97 ± 0.22 | 1.05 ± 0.09 |  |
|  | Intervention | 1.3 ± 0.15 | 1.5 ± 0.08 | 1.53 ± 0.27 | 1.4 ± 0.07 |  |
| Vegetables (excluding lettuce and white potatoes, cups/day) | Control (JN) | -0.08 ± 0.12 | 0.02 ± 0.05 | -0.18 ± 0.14 | -0.11 ± 0.06 | Group: 0.047 Sex: 0.1 Ethnicity: 0.156 Group x Sex: 0.991 Group x Ethnicity: 0.752 Sex x Ethnicity: 0.594 Group x Sex x Ethnicity: 0.728 |
|  | Intervention (JN) | 0.04 ± 0.1 | 0.22 ± 0.05 | -0.02 ± 0.16 | 0.05 ± 0.05 |  |
|  | Control | 0.46 ± 0.14 | 0.51 ± 0.06 | 0.38 ± 0.17 | 0.42 ± 0.08 |  |
|  | Intervention | 0.64 ± 0.12 | 0.86 ± 0.09 | 0.55 ± 0.18 | 0.6 ± 0.06 |  |
| Vegetable Soup (cups/day) | Control (JN) | -0.22 ± 0.12 | -0.09 ± 0.05 | -0.16 ± 0.15 | -0.1 ± 0.07 | Group: 0.031 Sex: 0.65 Ethnicity: 0.529 Group x Sex: 0.445 Group x Ethnicity: 0.496 Sex x Ethnicity: 0.271 Group x Sex x Ethnicity: 0.122 |
|  | Intervention (JN) | 0.18 ± 0.11 | -0.01 ± 0.05 | -0.09 ± 0.15 | 0.1 ± 0.05 |  |
|  | Control | 0.1 ± 0.05 | 0.14 ± 0.02 | 0.13 ± 0.06 | 0.2 ± 0.03 |  |
|  | Intervention | 0.18 ± 0.04 | 0.14 ± 0.02 | 0.13 ± 0.06 | 0.22 ± 0.02 |  |
| Lettuce (cups/day) | Control | 0.68 ± 0.11 | 0.57 ± 0.05 | 0.47 ± 0.13 | 0.55 ± 0.06 | Group: 0.213 Sex: 0.975 Ethnicity: 0.687 Group x Sex: 0.085 Group x Ethnicity: 0.885 Sex x Ethnicity: 0.771 Group x Sex x Ethnicity: 0.219 |
|  | Intervention | 0.59 ± 0.09 | 0.61 ± 0.04 | 0.76 ± 0.15 | 0.67 ± 0.04 |  |
| Cooked Dried Beans (cups/day) | Control (JN) | -0.33 ± 0.12^ab^ | -0.1 ± 0.05^ab^ | 0.03 ± 0.14^ab^ | -0.07 ± 0.06^a^ | Group: 0.222 Sex: 0.034 Ethnicity: 0.024 Group x Sex: 0.498 Group x Ethnicity: 0.173 Sex x Ethnicity: 0.921 Group x Sex x Ethnicity: 0.012 |
|  | Intervention (JN) | -0.1 ± 0.1^ab^ | -0.03 ± 0.04^a^ | -0.18 ± 0.15^ab^ | 0.25 ± 0.05^b^ |  |
|  | Control | 0.04 ± 0.03 | 0.06 ± 0.01 | 0.04 ± 0.04 | 0.1 ± 0.02 |  |
|  | Intervention | 0.06 ± 0.02 | 0.08 ± 0.01 | 0.05 ± 0.04 | 0.14 ± 0.01 |  |
| Tomato Sauce (cups/day) | Control (JN) | -0.23 ± 0.12 | -0.09 ± 0.05 | 0.04 ± 0.15 | 0.04 ± 0.07 | Group: 0.599 Sex: 0.002 Ethnicity: 0.31 Group x Sex: 0.607 Group x Ethnicity: 0.929 Sex x Ethnicity: 0.865 Group x Sex x Ethnicity: 0.453 |
|  | Intervention (JN) | -0.26 ± 0.11 | -0.22 ± 0.04 | -0.04 ± 0.17 | 0.09 ± 0.05 |  |
|  | Control | 0.07 ± 0.03 | 0.09 ± 0.01 | 0.09 ± 0.03 | 0.13 ± 0.02 |  |
|  | Intervention | 0.09 ± 0.02 | 0.09 ± 0.01 | 0.12 ± 0.04 | 0.16 ± 0.01 |  |
| Rice (grams/day) | Control (JN) | -0.34 ± 0.12 | -0.13 ± 0.04 | -0.02 ± 0.13 | 0.01 ± 0.05 | Group: 0.52 Sex: <0.001 Ethnicity: 0.093 Group x Sex: 0.298 Group x Ethnicity: 0.871 Sex x Ethnicity: 0.46 Group x Sex x Ethnicity: 0.486 |
|  | Intervention (JN) | -0.31 ± 0.09 | -0.2 ± 0.04 | 0.07 ± 0.14 | 0.16 ± 0.05 |  |
|  | Control | 33.38 ± 6.17 | 37.48 ± 2.29 | 45.88 ± 7 | 47.23 ± 3.04 |  |
|  | Intervention | 32.91 ± 5.39 | 35.44 ± 2.29 | 57.78 ± 8.82 | 51.99 ± 2.57 |  |
| Cereal (grams/day) | Control | 11.51 ± 2.27 | 15.81 ± 0.91 | 17.87 ± 2.91 | 18.66 ± 1.21 | Group: 0.79 Sex: 0.021 Ethnicity: 0.137 Group x Sex: 0.376 Group x Ethnicity: 0.776 Sex x Ethnicity: 0.944 Group x Sex x Ethnicity: 0.248 |
|  | Intervention | 15.28 ± 2.06 | 15.41 ± 0.77 | 15.81 ± 3.28 | 19.08 ± 0.99 |  |
| Estimated Fat Intake (% energy) | Control | 33.15 ± 0.46^a^ | 31.84 ± 0.19^ab^ | 31.42 ± 0.57^ab^ | 32.23 ± 0.24^ab^ | Group: 0.03 Sex: 0.615 Ethnicity: 0.923 Group x Sex: 0.003 Group x Ethnicity: 0.395 Sex x Ethnicity: 0.073 Group x Sex x Ethnicity: 0.046 |
|  | Intervention | 30.93 ± 0.36^b^ | 31.15 ± 0.16^ab^ | 31.91 ± 0.64^ab^ | 32.07 ± 0.17^ab^ |  |
| Processed Meat (grams/day) | Control (JN) | 0.11 ± 0.1 | -0.15 ± 0.04 | 0.21 ± 0.13 | 0.12 ± 0.06 | Group: 0.041 Sex: <0.001 Ethnicity: 0.113 Group x Sex: 0.152 Group x Ethnicity: 0.181 Sex x Ethnicity: 0.841 Group x Sex x Ethnicity: 0.194 |
|  | Intervention (JN) | -0.29 ± 0.09 | -0.24 ± 0.04 | 0.13 ± 0.13 | 0.05 ± 0.05 |  |
|  | Control | 12.16 ± 2.48 | 9 ± 0.93 | 17.5 ± 3.41 | 19.57 ± 1.68 |  |
|  | Intervention | 8.06 ± 2.01 | 8.46 ± 0.86 | 14.14 ± 3.33 | 18.31 ± 0.95 |  |
| Bacon or Sausage (grams/day) | Control (JN) | 0.2 ± 0.11^a^ | -0.04 ± 0.04^ab^ | 0.18 ± 0.13^ab^ | 0.11 ± 0.06^ab^ | Group: 0.343 Sex: <0.001 Ethnicity: 0.042 Group x Sex: 0.006 Group x Ethnicity: 0.717 Sex x Ethnicity: 0.375 Group x Sex x Ethnicity: 0.034 |
|  | Intervention (JN) | -0.22 ± 0.09^b^ | -0.13 ± 0.04^ab^ | 0.39 ± 0.15^a^ | 0.09 ± 0.04^ab^ |  |
|  | Control | 3.84 ± 0.58 | 2.57 ± 0.22 | 5.07 ± 0.75 | 3.81 ± 0.31 |  |
|  | Intervention | 2.03 ± 0.46 | 2.69 ± 0.21 | 5.64 ± 2.08 | 3.7 ± 0.22 |  |
| Hot Dogs (grams/day) | Control (JN) | 0.06 ± 0.11 | -0.2 ± 0.04 | 0.25 ± 0.14 | 0.11 ± 0.06 | Group: 0.017 Sex: <0.001 Ethnicity: 0.19 Group x Sex: 0.623 Group x Ethnicity: 0.082 Sex x Ethnicity: 0.458 Group x Sex x Ethnicity: 0.788 |
|  | Intervention (JN) | -0.28 ± 0.09 | -0.28 ± 0.04 | 0.01 ± 0.14 | 0.07 ± 0.04 |  |
|  | Control | 7.97 ± 2.26 | 6.06 ± 0.82 | 13.74 ± 3.51 | 15.78 ± 1.45 |  |
|  | Intervention | 5.9 ± 1.85 | 5.55 ± 0.79 | 8.18 ± 2.98 | 15.63 ± 1.02 |  |
| Cheese (grams/day) | Control (JN) | 0.05 ± 0.11 | -0.11 ± 0.04 | 0.08 ± 0.14 | 0.01 ± 0.06 | Group: 0.078 Sex: 0.044 Ethnicity: 0.631 Group x Sex: 0.371 Group x Ethnicity: 0.263 Sex x Ethnicity: 0.283 Group x Sex x Ethnicity: 0.096 |
|  | Intervention (JN) | -0.34 ± 0.1 | -0.11 ± 0.04 | 0.05 ± 0.16 | -0.1 ± 0.04 |  |
|  | Control | 12.04 ± 1.31 | 10.2 ± 0.55 | 12.22 ± 1.49 | 12.23 ± 0.71 |  |
|  | Intervention | 7.91 ± 1.05 | 10.25 ± 0.47 | 16.82 ± 4.3 | 11.1 ± 0.5 |  |
| Eggs (grams/day) | Control (JN) | 0.12 ± 0.11 | -0.02 ± 0.04 | 0.16 ± 0.14 | 0.12 ± 0.05 | Group: 0.147 Sex: 0.059 Ethnicity: 0.291 Group x Sex: 0.587 Group x Ethnicity: 0.76 Sex x Ethnicity: 0.175 Group x Sex x Ethnicity: 0.475 |
|  | Intervention (JN) | -0.01 ± 0.1 | -0.19 ± 0.04 | 0.01 ± 0.15 | 0.1 ± 0.05 |  |
|  | Control | 19.08 ± 2.72 | 18.11 ± 1.13 | 23.87 ± 4.22 | 20.37 ± 1.35 |  |
|  | Intervention | 19.27 ± 2.31 | 13.7 ± 0.95 | 21.25 ± 3.82 | 23.03 ± 1.19 |  |
| French Fries (grams/day) | Control (JN) | 0.12 ± 0.11 | -0.11 ± 0.04 | 0.11 ± 0.14 | 0.23 ± 0.05 | Group: 0.224 Sex: 0.009 Ethnicity: 0.965 Group x Sex: 0.891 Group x Ethnicity: 0.417 Sex x Ethnicity: 0.088 Group x Sex x Ethnicity: 0.277 |
|  | Intervention (JN) | -0.11 ± 0.09 | -0.1 ± 0.04 | 0.04 ± 0.15 | 0.13 ± 0.05 |  |
|  | Control | 27 ± 2.78 | 17.51 ± 0.95 | 23.31 ± 3.22 | 26.21 ± 1.43 |  |
|  | Intervention | 16.62 ± 2.19 | 18.52 ± 1.01 | 24.91 ± 3.64 | 23.5 ± 1.04 |  |
| Skim Milk (cups/day) | Control (JN) | -0.03 ± 0.1 | -0.1 ± 0.05 | 0.01 ± 0.13 | -0.1 ± 0.05 | Group: 0.301 Sex: 0.163 Ethnicity: 0.135 Group x Sex: 0.313 Group x Ethnicity: 0.996 Sex x Ethnicity: 0.196 Group x Sex x Ethnicity: 0.313 |
|  | Intervention (JN) | -0.07 ± 0.09 | -0.03 ± 0.04 | 0.21 ± 0.13 | -0.02 ± 0.04 |  |
|  | Control | 0.31 ± 0.07 | 0.33 ± 0.03 | 0.52 ± 0.1 | 0.41 ± 0.04 |  |
|  | Intervention | 0.29 ± 0.05 | 0.35 ± 0.02 | 0.41 ± 0.09 | 0.4 ± 0.02 |  |
| IPAQ Score | Control | 2258.41 ± 220.18^a^ | 2331.19 ± 93.27^a^ | 3060.29 ± 277.91^b^ | 2976.21 ± 121.22^b^ | Group: 0.964 Sex: 0.028 Ethnicity: 0.537 Group x Sex: 0.011 Group x Ethnicity: 0.475 Sex x Ethnicity: 0.483 Group x Sex x Ethnicity: 0.205 |
|  | Intervention | 2706.58 ± 191.74 | 2618.89 ± 89.79 | 2391.45 ± 409.16 | 2874.57 ± 103.06 |  |
| Vigorous Physical Activity (MET minutes/week) | Control | 994.39 ± 157.99^a^ | 1029.34 ± 61.39^a^ | 1514.62 ± 209.22^b^ | 1465.91 ± 86.75^b^ | Group: 0.825 Sex: 0.014 Ethnicity: 0.508 Group x Sex: 0.01 Group x Ethnicity: 0.462 Sex x Ethnicity: 0.356 Group x Sex x Ethnicity: 0.187 |
|  | Intervention | 1322.88 ± 130.24 | 1247.02 ± 61.54 | 1096.02 ± 211.08 | 1441.62 ± 64.91 |  |
| Moderate Physical Activity (MET Minutes/Week) | Control (JN) | 0.09 ± 0.1 | 0.14 ± 0.04 | 0.26 ± 0.13 | 0.35 ± 0.05 | Group: 0.815 Sex: 0.01 Ethnicity: 0.164 Group x Sex: 0.525 Group x Ethnicity: 0.824 Sex x Ethnicity: 0.382 Group x Sex x Ethnicity: 0.481 |
|  | Intervention (JN) | 0.17 ± 0.09 | 0.17 ± 0.04 | 0.19 ± 0.13 | 0.38 ± 0.04 |  |
|  | Control | 468.9 ± 71.13 | 472.42 ± 29.81 | 643.09 ± 114.24 | 605.98 ± 41.64 |  |
|  | Intervention | 440.86 ± 59.62 | 466.17 ± 27.36 | 414.22 ± 112.57 | 630.36 ± 31.8 |  |
| Walking Physical Activity (MET Mins/Week) | Control | 839.99 ± 90.17 | 866.97 ± 39.35 | 960.16 ± 113.3 | 954.15 ± 48.38 | Group: 0.487S Sex: 0.424 Ethnicity: 0.954 Group x Sex: 0.242 Group x Ethnicity: 0.891 Sex x Ethnicity: 0.591 Group x Sex x Ethnicity: 0.83 |
|  | Intervention | 848.15 ± 77.34 | 882.75 ± 37.6 | 870.43 ± 121.86 | 827.3 ± 35.93 |  |
| Total Stress Score | Control | 24.57 ± 0.78 | 25.9 ± 0.35 | 24.68 ± 0.96 | 23.82 ± 0.4 | Group: 0.955 Sex: 0.36 Ethnicity: 0.421 Group x Sex: 0.167 Group x Ethnicity: 0.747 Sex x Ethnicity: 0.157 Group x Sex x Ethnicity: 0.303 |
|  | Intervention | 24.26 ± 0.67 | 24.97 ± 0.31 | 24.65 ± 0.93 | 24.97 ± 0.3 |  |
| Positive Stress Subscale Score | Control | 19.38 ± 0.45^ab^ | 18.28 ± 0.2^a^ | 17.83 ± 0.61^ab^ | 19.21 ± 0.22^b^ | Group: 0.735 Sex: 0.400 Ethnicity: 0.826 Group x Sex: 0.752 Group x Ethnicity: 0.476 Sex x Ethnicity: 0.033 Group x Sex x Ethnicity: 0.011 |
|  | Intervention | 18.93 ± 0.42 | 18.78 ± 0.18 | 18.9 ± 0.56 | 18.52 ± 0.16 |  |
| Negative Stress Subscale Score | Control | 15.96 ± 0.53 | 16.19 ± 0.21 | 14.49 ± 0.61 | 15 ± 0.26 | Group: 0.906 Sex: 0.008 Ethnicity: 0.134 Group x Sex: 0.059 Group x Ethnicity: 0.713 Sex x Ethnicity: 0.785 Group x Sex x Ethnicity: 0.447 |
|  | Intervention | 15.13 ± 0.4 | 16.02 ± 0.2 | 15.19 ± 0.66 | 15.47 ± 0.22 |  |
| Nightly Sleep Duration (hours) | Control | 6.96 ± 0.19 | 7.2 ± 0.07 | 7.19 ± 0.2 | 7.18 ± 0.09 | Group: 0.037 Sex: 0.978 Ethnicity: 0.115 Group x Sex: 0.376 Group x Ethnicity: 0.619 Sex x Ethnicity: 0.708 Group x Sex x Ethnicity: 0.434 |
|  | Intervention | 7.34 ± 0.15 | 7.52 ± 0.07 | 7.2 ± 0.26 | 7.46 ± 0.07 |  |

Post-study data are presented as model-adjusted least squares means ± SE. JN indicates JN transformed variables. Models were fit using baseline-adjusted linear regression in the combined sample from two randomized controlled trials. Fixed effects included Group, Sex, and Ethnicity, along with all two- and three-way interaction terms. All models adjusted for the baseline value of the outcome, baseline BMI, study, and state. Missing outcome data were addressed using multiple imputation by chained equations with predictive mean matching. Analyses were conducted within each imputed dataset, and estimates were combined using Rubin’s rules. Joint tests represent omnibus tests of each model term (e.g., Group, Sex, Ethnicity, and interaction terms) derived from pooled model estimates using the emmeans framework. Pairwise comparisons were calculated from the pooled estimates, with p-values adjusted using the multivariate t method.

P-values correspond to transformed variables unless transformation was not required to meet model assumptions, in which case non-transformed values are reported. Non-transformed data are presented for interpretability only when transformations were applied for analysis.

Superscript letters indicate pairwise differences among subgroups within significant Group x Sex, Group x Ethnicity and Group x Sex x Ethnicity interaction effects. When both lower- and higher-order interaction terms involving the same factors were significant (e.g., Group × Sex and Group × Sex × Ethnicity), pairwise comparisons were interpreted for the highest-order interaction. Values sharing at least one letter are not significantly different.

BL; baseline, JN; Johnson’s Normalization transformation

**Supplementary Table 4. Model-Adjusted Group, Sex, and Race Effects on Post-Study Outcomes in Young College Adults**

| **Outcome** | **Group** | **Female Black** | **Female White** | **Female Other** | **Male Black** | **Male White** | **Male Other** | **BL-adjusted P-values** |
| --- | --- | --- | --- | --- | --- | --- | --- | --- |
| Weight (kg) | Control | 70.13 ± 0.45 | 70.06 ± 0.19 | 69.55 ± 0.38 | 69.59 ± 0.65 | 70.37 ± 0.3 | 70.56 ± 0.46 | Group: 0.147  Sex: 0.185  Race: 0.895  Group x Sex: 0.504  Group x Race: 0.372  Sex x Race: 0.125  Group x Sex x Race: 0.878 |
|  | Intervention | 70.9 ± 1.51 | 69.94 ± 0.2 | 69.52 ± 0.42 | 71.11 ± 0.63 | 70.36 ± 0.29 | 71.01 ± 0.54 |  |
| Waist Circumference (cm) | Control | 80.11 ± 0.52 | 81.23 ± 0.21 | 80.98 ± 0.43 | 80.77 ± 0.71 | 82.17 ± 0.3 | 82.46 ± 0.51 | Group: 0.722  Sex: 0.031  Race: 0.071  Group x Sex: 0.282  Group x Race: 0.897  Sex x Race: 0.326  Group x Sex x Race: 0.767 |
|  | Intervention | 81.19 ± 1.09 | 81.36 ± 0.2 | 81.15 ± 0.46 | 80.42 ± 0.75 | 82.02 ± 0.31 | 82.28 ± 0.65 |  |
| NCIFV Score | Control (JN) | -0.23 ± 0.11 | -0.11 ± 0.05 | -0.19 ± 0.09 | -0.19 ± 0.16 | -0.17 ± 0.06 | -0.19 ± 0.12 | Group: <0.001  Sex: 0.142  Race: 0.968  Group x Sex: 0.138  Group x Race: 0.522  Sex x Race: 0.194  Group x Sex x Race: 0.468 |
|  | Intervention (JN) | 0.08 ± 0.1 | 0.03 ± 0.04 | -0.12 ± 0.1 | 0.19 ± 0.16 | 0.06 ± 0.08 | 0.27 ± 0.13 |  |
|  | Control | 2.78 ± 0.28 | 2.92 ± 0.11 | 2.93 ± 0.24 | 2.94 ± 0.38 | 2.81 ± 0.16 | 2.77 ± 0.31 |  |
|  | Intervention | 3.76 ± 0.74 | 3.19 ± 0.11 | 3.16 ± 0.25 | 2.83 ± 0.44 | 3.42 ± 0.41 | 3.98 ± 0.32 |  |
| Fruit and Fruit Juice (cups/day) | Control (JN) | -0.35 ± 0.1 | -0.11 ± 0.04 | -0.2 ± 0.09 | -0.18 ± 0.15 | -0.13 ± 0.06 | -0.08 ± 0.12 | Group: 0.003  Sex: 0.057  Race: 0.417  Group x Sex: 0.706  Group x Race: 0.255  Sex x Race: 0.307  Group x Sex x Race: 0.753 |
|  | Intervention (JN) | 0.03 ± 0.09 | 0.03 ± 0.04 | -0.19 ± 0.09 | 0.11 ± 0.16 | 0.08 ± 0.09 | 0.06 ± 0.12 |  |
|  | Control | 0.88 ± 0.14 | 1.1 ± 0.06 | 1.1 ± 0.12 | 1.02 ± 0.2 | 1.12 ± 0.08 | 1.25 ± 0.17 |  |
|  | Intervention | 1.35 ± 0.14 | 1.27 ± 0.05 | 1.18 ± 0.11 | 1.47 ± 0.23 | 1.34 ± 0.11 | 1.44 ± 0.15 |  |
| Fruit (cups/day) | Control (JN) | -0.21 ± 0.1 | 0 ± 0.04^*^ | -0.21 ± 0.09 | -0.25 ± 0.15 | -0.19 ± 0.06 | -0.12 ± 0.11 | Group: <0.001  Sex: 0.939  Race: 0.537  Group x Sex: 0.441  Group x Race: 0.13  Sex x Race: 0.033  Group x Sex x Race: 0.825 |
|  | Intervention (JN) | 0.11 ± 0.1 | 0.1 ± 0.04^*^ | -0.05 ± 0.09 | 0.27 ± 0.16 | -0.02 ± 0.07 | 0.03 ± 0.11 |  |
|  | Control | 0.55 ± 0.1 | 0.68 ± 0.04 | 0.55 ± 0.08 | 0.53 ± 0.14 | 0.52 ± 0.05 | 0.64 ± 0.11 |  |
|  | Intervention | 0.85 ± 0.1 | 0.73 ± 0.04 | 0.69 ± 0.08 | 0.87 ± 0.14 | 0.69 ± 0.11 | 0.66 ± 0.09 |  |
| All Vegetables (cups/day) | Control (JN) | -0.18 ± 0.11 | -0.03 ± 0.04 | -0.02 ± 0.09 | -0.1 ± 0.15 | -0.18 ± 0.06 | -0.17 ± 0.11 | Group: 0.001  Sex: 0.728  Race: 0.571  Group x Sex: 0.093  Group x Race: 0.68  Sex x Race: 0.341  Group x Sex x Race: 0.284 |
|  | Intervention (JN) | 0.07 ± 0.09 | 0.06 ± 0.04 | 0.03 ± 0.09 | 0.09 ± 0.17 | 0.06 ± 0.08 | 0.33 ± 0.13 |  |
|  | Control | 1.12 ± 0.14 | 1.17 ± 0.06 | 1.31 ± 0.13 | 1.42 ± 0.19 | 1.03 ± 0.08 | 1.13 ± 0.16 |  |
|  | Intervention | 1.5 ± 0.18 | 1.29 ± 0.06 | 1.31 ± 0.13 | 1.11 ± 0.2 | 1.35 ± 0.09 | 1.81 ± 0.19 |  |
| Vegetables (excluding lettuce and white potatoes, cups/day) | Control (JN) | -0.08 ± 0.11 | 0.07 ± 0.04 | -0.21 ± 0.09 | 0.02 ± 0.14 | -0.13 ± 0.06 | -0.1 ± 0.13 | Group: 0.006  Sex: 0.593  Race: 0.777  Group x Sex: 0.515  Group x Race: 0.158  Sex x Race: 0.354  Group x Sex x Race: 0.152 |
|  | Intervention (JN) | 0.12 ± 0.09 | 0.12 ± 0.04 | 0.16 ± 0.09 | -0.09 ± 0.15 | 0.09 ± 0.08 | 0.19 ± 0.13 |  |
|  | Control | 0.52 ± 0.09 | 0.56 ± 0.04 | 0.56 ± 0.09 | 0.6 ± 0.12 | 0.41 ± 0.06 | 0.59 ± 0.11 |  |
|  | Intervention | 0.6 ± 0.08 | 0.6 ± 0.04 | 0.61 ± 0.08 | 0.4 ± 0.12 | 0.6 ± 0.07 | 0.79 ± 0.15 |  |
| Vegetable Soup (cups/day) | Control (JN) | -0.33 ± 0.11 | -0.01 ± 0.05 | -0.13 ± 0.09 | -0.02 ± 0.16 | -0.13 ± 0.07 | -0.05 ± 0.13 | Group: 0.258  Sex: 0.347  Race: 0.504  Group x Sex: 0.618  Group x Race: 0.98  Sex x Race: 0.071  Group x Sex x Race: 0.273 |
|  | Intervention (JN) | -0.19 ± 0.09 | 0 ± 0.04 | 0.05 ± 0.1 | 0.04 ± 0.16 | 0.01 ± 0.09 | -0.11 ± 0.13 |  |
|  | Control | 0.13 ± 0.04 | 0.15 ± 0.02 | 0.12 ± 0.03 | 0.32 ± 0.09 | 0.15 ± 0.02 | 0.3 ± 0.07^b^ |  |
|  | Intervention | 0.12 ± 0.03 | 0.15 ± 0.02 | 0.15 ± 0.03 | 0.16 ± 0.05 | 0.14 ± 0.02 | 0.18 ± 0.04 |  |
| Lettuce (cups/day) | Control (JN) | -0.25 ± 0.09 | -0.04 ± 0.04 | -0.01 ± 0.08 | -0.12 ± 0.14 | -0.14 ± 0.06 | -0.17 ± 0.1 | Group: 0.001  Sex: 0.412  Race: 0.502  Group x Sex: 0.092  Group x Race: 0.543  Sex x Race: 0.456  Group x Sex x Race: 0.154 |
|  | Intervention (JN) | 0.02 ± 0.1 | 0.03 ± 0.04 | -0.02 ± 0.09 | 0.06 ± 0.16 | 0.06 ± 0.07 | 0.3 ± 0.1 |  |
|  | Control | 0.45 ± 0.09 | 0.54 ± 0.04 | 0.62 ± 0.09 | 0.65 ± 0.13 | 0.51 ± 0.06 | 0.46 ± 0.1 |  |
|  | Intervention | 0.76 ± 0.11 | 0.6 ± 0.04 | 0.68 ± 0.09 | 0.5 ± 0.14 | 0.64 ± 0.09 | 1.01 ± 0.12 |  |
| Cooked Dried Beans (cups/day) | Control (JN) | -0.11 ± 0.11 | -0.11 ± 0.05 | -0.09 ± 0.09 | 0.13 ± 0.15 | -0.07 ± 0.06 | -0.12 ± 0.11 | Group: 0.179  Sex: 0.016  Race: 0.932  Group x Sex: 0.359  Group x Race: 0.23  Sex x Race: 0.346  Group x Sex x Race: 0.345 |
|  | Intervention (JN) | -0.17 ± 0.09 | 0.01 ± 0.05 | -0.06 ± 0.09 | 0.05 ± 0.16 | 0.06 ± 0.1 | 0.25 ± 0.12 |  |
|  | Control | 0.07 ± 0.03 | 0.06 ± 0.01 | 0.06 ± 0.02 | 0.13 ± 0.04 | 0.1 ± 0.02 | 0.06 ± 0.03 |  |
|  | Intervention | 0.07 ± 0.02 | 0.07 ± 0.01 | 0.12 ± 0.02 | 0.08 ± 0.04 | 0.12 ± 0.06 | 0.14 ± 0.03 |  |
| Tomato Sauce (cups/day) | Control (JN) | -0.2 ± 0.11 | -0.15 ± 0.05 | -0.1 ± 0.09 | -0.04 ± 0.18 | 0.11 ± 0.07 | -0.22 ± 0.12 | Group: 0.589  Sex: 0.013  Race: 0.531  Group x Sex: 0.304  Group x Race: 0.317  Sex x Race: 0.391  Group x Sex x Race: 0.125 |
|  | Intervention (JN) | -0.06 ± 0.12 | -0.21 ± 0.04 | -0.25 ± 0.1 | 0.07 ± 0.18 | 0.03 ± 0.08 | 0.07 ± 0.11 |  |
|  | Control | 0.08 ± 0.02 | 0.08 ± 0.01 | 0.09 ± 0.02 | 0.18 ± 0.04 | 0.12 ± 0.01 | 0.09 ± 0.02 |  |
|  | Intervention | 0.12 ± 0.02 | 0.08 ± 0.01 | 0.11 ± 0.02 | 0.14 ± 0.04 | 0.16 ± 0.05 | 0.14 ± 0.03 |  |
|  | Control (JN) | -0.11 ± 0.1^a^ | -0.2 ± 0.04^a^ | -0.1 ± 0.09^a^ | -0.07 ± 0.13^a^ | 0.04 ± 0.06^a^ | -0.11 ± 0.12^a^ | Group: 0.037  Sex: <0.001  Race: 0.311  Group x Sex: 0.015  Group x Race: 0.179  Sex x Race: 0.502  Group x Sex x Race: 0.065 |
| Rice (grams/day) | Intervention (JN) | -0.11 ± 0.09^a^ | -0.19 ± 0.04^a^ | -0.13 ± 0.08^a^ | 0.08 ± 0.13^b^ | 0.09 ± 0.07^b^ | 0.43 ± 0.11^b^ |  |
|  | Control | 41.29 ± 4.9 | 35.07 ± 2.12 | 38.15 ± 4.61 | 41.79 ± 6.93 | 46.77 ± 3.04 | 49.79 ± 6.52 |  |
|  | Intervention | 46.88 ± 4.95 | 33.46 ± 1.98 | 36.57 ± 4.41 | 43.83 ± 7.3 | 49.27 ± 3.09 | 82.84 ± 7.48 |  |
| Cereal (grams/day) | Control (JN) | -0.19 ± 0.1 | -0.07 ± 0.05 | -0.21 ± 0.09 | 0.16 ± 0.14 | 0 ± 0.06 | -0.06 ± 0.11 | Group: 0.526  Sex: 0.156  Race: 0.732  Group x Sex: 0.073  Group x Race: 0.619  Sex x Race: 0.544  Group x Sex x Race: 0.579 |
|  | Intervention (JN) | -0.07 ± 0.11 | -0.07 ± 0.04 | -0.13 ± 0.09 | -0.13 ± 0.15 | -0.15 ± 0.11 | -0.07 ± 0.13 |  |
|  | Control | 15.17 ± 2.11 | 15.41 ± 0.91 | 14.22 ± 1.71 | 22.29 ± 3.43 | 17.35 ± 1.32 | 21.63 ± 2.7 |  |
|  | Intervention | 18.71 ± 2.44 | 15.72 ± 0.85 | 14.6 ± 1.82 | 15.35 ± 3.27 | 17.87 ± 1.44 | 20.31 ± 3.02 |  |
| Estimated Fat Intake (% energy) | Control (JN) | 0.08 ± 0.09 | 0 ± 0.04 | 0.15 ± 0.08 | 0.01 ± 0.13 | 0.14 ± 0.06 | 0.18 ± 0.1 | Group: 0.188  Sex: 0.422  Race: 0.75  Group x Sex: 0.799  Group x Race: 0.109  Sex x Race: 0.087  Group x Sex x Race: 0.165 |
|  | Intervention (JN) | 0.01 ± 0.08 | -0.1 ± 0.04 | 0.04 ± 0.08 | 0.23 ± 0.16 | 0.06 ± 0.11 | -0.18 ± 0.12 |  |
|  | Control | 31.86 ± 0.44 | 31.66 ± 0.18 | 32.49 ± 0.34 | 31.71 ± 0.59 | 32.2 ± 0.24 | 32.24 ± 0.47 |  |
|  | Intervention | 31.79 ± 0.4 | 31.18 ± 0.17 | 31.75 ± 0.37 | 32.57 ± 0.76 | 32.03 ± 0.59 | 30.47 ± 0.47 |  |
| Processed Meat (grams/day) | Control (JN) | -0.1 ± 0.09 | -0.21 ± 0.04 | -0.08 ± 0.08 | 0.03 ± 0.15 | 0.12 ± 0.06 | 0.05 ± 0.12 | Group: 0.428  Sex: <0.001  Race: 0.027  Group x Sex: 0.06  Group x Race: 0.011  Sex x Race: 0.263  Group x Sex x Race: 0.134 |
|  | Intervention (JN) | -0.04 ± 0.09^a^ | -0.26 ± 0.04^b^ | -0.25 ± 0.08^b^ | 0.58 ± 0.13^a^ | 0.1 ± 0.07^b^ | -0.04 ± 0.12^b^ |  |
|  | Control | 11.03 ± 2.11 | 7.21 ± 0.96 | 14.04 ± 1.97 | 16.1 ± 3.21 | 18.97 ± 1.8 | 21.93 ± 3.3 |  |
|  | Intervention | 11.71 ± 1.99 | 7.39 ± 0.89 | 10.31 ± 1.9 | 33.22 ± 4.37 | 16.41 ± 1.5 | 19.35 ± 2.75 |  |
| Bacon or Sausage (grams/day) | Control (JN) | 0 ± 0.1 | -0.08 ± 0.04^*^ | 0 ± 0.08 | 0.12 ± 0.14^‖^ | 0.13 ± 0.06^‡^ | -0.1 ± 0.11 | Group: 0.638  Sex: 0.001  Race: <0.001  Group x Sex: 0.08  Group x Race: 0.013  Sex x Race: 0.023  Group x Sex x Race: 0.698 |
|  | Intervention (JN) | 0.11 ± 0.08^a^ | -0.13 ± 0.04^b,*^ | -0.25 ± 0.09^c^ | 0.53 ± 0.13^a,‖^ | 0.17 ± 0.06^b,‡^ | -0.19 ± 0.11^c^ |  |
|  | Control | 3.19 ± 0.5 | 2.55 ± 0.22 | 2.97 ± 0.43 | 4.41 ± 0.67 | 3.85 ± 0.33 | 3.85 ± 0.68 |  |
|  | Intervention | 3.92 ± 0.49 | 2.4 ± 0.21 | 2.29 ± 0.45 | 6.55 ± 1.79 | 4.05 ± 0.32 | 2.77 ± 0.62 |  |
| Hot Dogs (grams/day) | Control (JN)^¶^ | -0.14 ± 0.1 | -0.29 ± 0.05 | -0.03 ± 0.08 | 0.13 ± 0.15 | 0.1 ± 0.06 | 0.13 ± 0.1 | Group: 0.517  Sex: <0.001  Race: 0.005  Group x Sex: 0.127  Group x Race: 0.198  Sex x Race: 0.398  Group x Sex x Race: 0.309 |
|  | Intervention (JN) ^¶^ | -0.11 ± 0.09 | -0.31 ± 0.04 | -0.17 ± 0.09 | 0.48 ± 0.14 | 0.06 ± 0.06 | 0.17 ± 0.13 |  |
|  | Control | 7.63 ± 2.12 | 4.61 ± 0.92 | 11.22 ± 1.88 | 12.63 ± 3.01 | 15.31 ± 1.61 | 19.88 ± 3.73 |  |
|  | Intervention | 7.94 ± 1.84 | 4.96 ± 0.85 | 8.05 ± 1.76 | 26.8 ± 3.83 | 13.04 ± 2.71 | 18.59 ± 3.17 |  |
| Cheese (grams/day) ^‡^ | Control | 9.14 ± 1.07 | 10.93 ± 0.53 | 9.15 ± 0.98 | 10.53 ± 1.62 | 13.11 ± 0.69 | 10.58 ± 1.27 | Group: 0.877  Sex: 0.003  Race: 0.018  Group x Sex: 0.687  Group x Race: 0.308  Sex x Race: 0.771  Group x Sex x Race: 0.75 |
|  | Intervention | 9.81 ± 1.32 | 10.37 ± 0.46 | 8.63 ± 0.94 | 13.32 ± 1.73 | 12.35 ± 1.74 | 9.61 ± 1.21 |  |
| Eggs (grams/day) | Control | 16.89 ± 2.31 | 16.03 ± 1.02^*^ | 19.31 ± 2.1 | 21.46 ± 3.11 | 20.01 ± 1.41 | 21.5 ± 2.66 | Group: 0.111  Sex: 0.002  Race: 0.368  Group x Sex: 0.674  Group x Race: 0.006  Sex x Race: 0.044  Group x Sex x Race: 0.068 |
|  | Intervention | 15.37 ± 2.05 | 14.4 ± 0.92^*^ | 14.29 ± 2.03 | 15.54 ± 3.48 | 26.05 ± 1.53 | 16.35 ± 2.73 |  |
| French Fries (grams/day) | Control (JN) | -0.02 ± 0.1 | -0.12 ± 0.04^*^ | -0.09 ± 0.08 | -0.12 ± 0.15 | 0.24 ± 0.06 | 0.3 ± 0.12 | Group: 0.993  Sex: 0.014  Race: 0.628  Group x Sex: 0.276  Group x Race: 0.477  Sex x Race: 0.039  Group x Sex x Race: 0.594 |
|  | Intervention (JN) | 0.06 ± 0.09 | -0.08 ± 0.04^*^ | -0.02 ± 0.08 | 0.01 ± 0.19 | 0.14 ± 0.06 | 0.07 ± 0.14 |  |
|  | Control | 20.63 ± 2.52 | 17.13 ± 1 | 20.6 ± 1.96 | 23.01 ± 4.25 | 25.18 ± 1.56 | 28.59 ± 2.68 |  |
|  | Intervention | 21.99 ± 2.36 | 17.49 ± 0.94 | 20.4 ± 2.02 | 25.96 ± 4.28 | 23.73 ± 1.46 | 28.06 ± 3.09 |  |
| Skim Milk (cups/day) | Control (JN) | -0.25 ± 0.1^¶^ | -0.08 ± 0.04 | -0.14 ± 0.08 | -0.2 ± 0.13 | -0.09 ± 0.06 | 0.05 ± 0.11 | Group: 0.149  Sex: 0.003  Race: 0.459  Group x Sex: 0.121  Group x Race: 0.266  Sex x Race: 0.015  Group x Sex x Race: 0.159 |
|  | Intervention (JN) | -0.28 ± 0.09^¶^ | 0 ± 0.04 | -0.18 ± 0.08 | 0.23 ± 0.14 | -0.01 ± 0.06 | 0.03 ± 0.11 |  |
|  | Control | 0.29 ± 0.06 | 0.33 ± 0.03 | 0.27 ± 0.05 | 0.37 ± 0.08 | 0.4 ± 0.04 | 0.54 ± 0.07 |  |
|  | Intervention | 0.24 ± 0.05 | 0.36 ± 0.02 | 0.27 ± 0.05 | 0.43 ± 0.09 | 0.42 ± 0.04 | 0.48 ± 0.07 |  |
| IPAQ Score | Control | 2169 ± 203.69 | 2388.75 ± 88.11 | 2341.79 ± 202.08 | 2408.2 ± 259.95 | 3095.22 ± 137.61 | 2945.31 ± 222.1 | Group: 0.452  Sex: <0.001  Race: 0.253  Group x Sex: 0.503  Group x Race: 0.416  Sex x Race: 0.605  Group x Sex x Race: 0.519 |
|  | Intervention | 2463.65 ± 212.36 | 2684.95 ± 86.74 | 2298.69 ± 171.49 | 2776.15 ± 291.22 | 2843.84 ± 354.32 | 2874.03 ± 207.75 |  |
| Vigorous Physical Activity (MET minutes/week) | Control | 1001.43 ± 142.68 | 1078.2 ± 63.86 | 951.44 ± 123.13 | 1301.96 ± 205.63 | 1531.02 ± 90.1 | 1367.09 ± 151.71 | Group: 0.552  Sex: <0.001  Race: 0.194  Group x Sex: 0.442  Group x Race: 0.679  Sex x Race: 0.973  Group x Sex x Race: 0.83 |
|  | Intervention | 1137.66 ± 125.93 | 1260.86 ± 60.25 | 974.22 ± 123.52 | 1474.13 ± 209.33 | 1434.73 ± 293 | 1269.84 ± 158.24 |  |
| Moderate Physical Activity (MET Minutes/Week) | Control (JN) ^¶^ | -0.08 ± 0.09 | 0.2 ± 0.04 | -0.01 ± 0.08 | 0.1 ± 0.12 | 0.36 ± 0.06 | 0.31 ± 0.1 | Group: 0.05  Sex: <0.001  Race: 0.005  Group x Sex: 0.883  Group x Race: 0.312  Sex x Race: 0.089  Group x Sex x Race: 0.856 |
|  | Intervention (JN) ^¶^ | 0.09 ± 0.08 | 0.26 ± 0.04 | 0.09 ± 0.08 | 0.28 ± 0.12 | 0.35 ± 0.06 | 0.43 ± 0.1 |  |
|  | Control | 374.33 ± 67.3 | 496.52 ± 29.93 | 437.22 ± 60.06 | 521.73 ± 98.17 | 629.41 ± 44.07 | 575.22 ± 78.09 |  |
|  | Intervention | 424.25 ± 57.04 | 519.36 ± 29.76 | 381.34 ± 53.43 | 449.24 ± 93.64 | 577.55 ± 48.37 | 682.03 ± 71.62 |  |
| Walking Physical Activity (MET Mins/Week) | Control | 765.82 ± 81.28 | 890.45 ± 38.37 | 862.73 ± 75.93 | 678.55 ± 115.17 | 993.14 ± 51.18 | 998.89 ± 97.66 | Group: 0.957  Sex: 0.574  Race: 0.098  Group x Sex: 0.706  Group x Race: 0.55  Sex x Race: 0.406  Group x Sex x Race: 0.793 |
|  | Intervention | 825.02 ± 78.7 | 907 ± 35.9 | 852.06 ± 73.92 | 798.5 ± 120.32 | 879.57 ± 174.41 | 944.16 ± 87.01 |  |
| Total Stress Score | Control | 25.98 ± 0.65 | 25.67 ± 0.32 | 26.05 ± 0.58 | 25.27 ± 1.13 | 24.18 ± 0.42 | 23.66 ± 0.73 | Group: 0.588  Sex: 0.02  Race: 0.405  Group x Sex: 0.391  Group x Race: 0.988  Sex x Race: 0.882  Group x Sex x Race: 0.567 |
|  | Intervention | 26.04 ± 0.74 | 24.87 ± 0.29 | 24.91 ± 0.61 | 24.81 ± 1.15 | 24.3 ± 1.55 | 24.41 ± 0.78 |  |
| Positive Stress Subscale Score | Control | 18.05 ± 0.4 | 18.58 ± 0.18 | 18.07 ± 0.36 | 18.68 ± 0.64 | 19.22 ± 0.24 | 18.8 ± 0.49 | Group: 0.608  Sex: 0.027  Race: 0.172  Group x Sex: 0.482  Group x Race: 0.816  Sex x Race: 0.662  Group x Sex x Race: 0.753 |
|  | Intervention | 18.33 ± 0.42 | 18.92 ± 0.17 | 18.35 ± 0.35 | 18.55 ± 0.64 | 18.92 ± 0.26 | 19.12 ± 0.44 |  |
| Negative Stress Subscale Score | Control | 16.12 ± 0.47 | 16.26 ± 0.2 | 16.35 ± 0.43 | 15.94 ± 0.69 | 15.15 ± 0.28 | 14.05 ± 0.46 | Group: 0.723  Sex: <0.001  Race: 0.194  Group x Sex: 0.441  Group x Race: 0.769  Sex x Race: 0.643  Group x Sex x Race: 0.103 |
|  | Intervention | 16.39 ± 0.39 | 15.89 ± 0.18 | 15.53 ± 0.4 | 15.23 ± 0.67 | 15.02 ± 0.69 | 15.17 ± 0.49 |  |
| Nightly Sleep Duration (hours) | Control | 6.69 ± 0.17 | 7.35 ± 0.08 | 6.95 ± 0.15 | 6.88 ± 0.2 | 7.22 ± 0.09 | 7.14 ± 0.16 | Group: 0.004  Sex: 0.899  Race: 0.18  Group x Sex: 0.516  Group x Race: 0.056  Sex x Race: 0.531  Group x Sex x Race: 0.428 |
|  | Intervention | 7.41 ± 0.15 | 7.45 ± 0.06 | 7.51 ± 0.37 | 7.6 ± 0.27 | 7.46 ± 0.25 | 7.15 ± 0.17 |  |

Post-study data are presented as model-adjusted least squares means ± SE. JN indicates JN transformed variables. Models were fit using baseline-adjusted linear regression in the combined sample from two randomized controlled trials. Fixed effects included Group, Sex, and Race, along with all two- and three-way interaction terms. All models adjusted for the baseline value of the outcome, baseline BMI, study, and state. Missing outcome data were addressed using multiple imputation by chained equations with predictive mean matching. Analyses were conducted within each imputed dataset, and estimates were combined using Rubin’s rules. Joint tests represent omnibus tests of each model term (e.g., Group, Sex, Race, and interaction terms) derived from pooled model estimates using the emmeans framework. Pairwise comparisons were calculated from the pooled estimates, with p-values adjusted using the multivariate t method.

P-values correspond to transformed variables unless transformation was not required to meet model assumptions, in which case non-transformed values are reported. Non-transformed data are presented for interpretability only when transformations were applied for analysis.

Superscript letters indicate pairwise differences among subgroups within significant Group x Sex, Group x Race and Group x Sex x Race interaction effects. When both lower- and higher-order interaction terms involving the same factors were significant (e.g., Group × Sex and Group × Sex × Race), pairwise comparisons were interpreted for the highest-order interaction. Values sharing at least one letter are not significantly different. ^*^ indicates Female vs. Male; ¶ indicates Black vs. White; ‖ indicates Black vs. Other; ‡ indicates White vs. Other, within subgroup when shown in subgroup columns and for overall sex (for ^*^) or race (for ¶, ‖, ‡) effects when shown outside subgroup columns.

BL; baseline, JN; Johnson’s Normalization transformation

**Supplemental Table 5. Model-Adjusted Significant Overall Group, Sex and Ethnicity Effects for Post-Study Outcomes in Young College Adults**

| **Outcome** | **Group:**  **Control**  **vs.**  **Intervention** | **Sex:**  **Female**  **vs.**  **Male** | **Ethnicity:**  **Hispanic**  **vs.**  **Non-Hispanic** |
| --- | --- | --- | --- |
| Fruit and Fruit Juice (cups/day)^**^ | 1.13 ± 0.08 1.42 ± 0.08 | 1.26 ± 0.06  1.29 ± 0.08 | 1.3 ± 0.09  1.25 ± 0.04 |
| Fruit (cups/day) | 0.57 ± 0.06  0.79 ± 0.06 | 0.76 ± 0.05  0.6 ± 0.06 | 0.7 ± 0.07  0.66 ± 0.03 |
| Fruit (cups/day) (JN)^**^ | -0.14 ± 0.06  0.02 ± 0.06 | 0 ± 0.04  -0.11 ± 0.06 | -0.08 ± 0.07  -0.04 ± 0.03 |
| All Vegetables (cups/day) | 1.11 ± 0.08  1.43 ± 0.09 | 1.31 ± 0.06  1.24 ± 0.09 | 1.26 ± 0.11  1.28 ± 0.04 |
| All Vegetables (cups/day) (JN)^*^ | -0.11 ± 0.05  0.03 ± 0.06 | 0.03 ± 0.04  -0.11 ± 0.06 | -0.1 ± 0.07  0.02 ± 0.03 |
| Vegetables (excluding lettuce and white potatoes, cups/day) | 0.44 ± 0.07  0.66 ± 0.07 | 0.62 ± 0.05  0.49 ± 0.07 | 0.51 ± 0.08  0.6 ± 0.03 |
| Vegetables (excluding lettuce and white potatoes, cups/day) (JN)^**^ | -0.09 ± 0.05  0.07 ± 0.06 | 0.05 ± 0.04  -0.06 ± 0.06 | -0.06 ± 0.07  0.05 ± 0.03 |
| Vegetable Soup (cups/day) | 0.14 ± 0.02  0.17 ± 0.02 | 0.14 ± 0.02  0.17 ± 0.02 | 0.13 ± 0.03  0.18 ± 0.01 |
| Vegetable Soup (cups/day) (JN)^**^ | -0.14 ± 0.06  0.05 ± 0.06 | -0.03 ± 0.04  0.61 ± 0.04 | -0.07 ± 0.07  -0.02 ± 0.03 |
| Tomato Sauce (cups/day) | 0.09 ± 0.01  0.11 ± 0.01 | 0.08 ± 0.01  0.12 ± 0.01 | 0.09 ± 0.01  0.12 ± 0.01 |
| Tomato Sauce (cups/day) (JN)^*^ | -0.06 ± 0.05  -0.11 ± 0.06 | -0.2 ± 0.05  0.03 ± 0.06 | -0.12 ± 0.07  -0.04 ± 0.03 |
| Rice (grams/day) | 40.99 ± 2.62  44.53 ± 3.32 | 34.8 ± 2.23  50.72 ± 3.2 | 42.49 ± 3.56  43.04 ± 1.59 |
| Rice (grams/day) (JN)^*^ | -0.12 ± 0.05  -0.07 ± 0.06 | -0.25 ± 0.04  0.06 ± 0.05 | -0.15 ± 0.06  -0.04 ± 0.03 |
| Cereal (grams/day)^*^ | 15.96 ± 1.05  16.4 ± 1.22 | 14.5 ± 0.82  17.86 ± 1.27 | 15.12 ± 1.38  17.24 ± 0.61 |
| Processed Meat (grams/day) | 14.56 ± 1.27  12.24 ± 1.15 | 9.42 ± 0.9  17.38 ± 1.32 | 12.96 ± 1.51  13.84 ± 0.66 |
| Processed Meat (grams/day) (JN)^*, **^ | 0.07 ± 0.05  -0.09 ± 0.05 | -0.14 ± 0.04  0.13 ± 0.05 | 0.04 ± 0.06  -0.05 ± 0.03 |
| Hot Dogs (grams/day) | 10.89 ± 1.2  8.82 ± 1.13 | 6.37 ± 0.82  13.33 ± 1.29 | 8.95 ± 1.39  10.75 ± 0.66 |
| Hot Dogs (grams/day) (JN)^*, **^ | 0.05 ± 0.05  -0.12 ± 0.05 | -0.18 ± 0.04  0.11 ± 0.05 | 0.01 ± 0.06  -0.08 ± 0.03 |
| Cheese (grams/day) | 11.67 ± 0.6  11.52 ± 1.13 | 10.1 ± 0.46  13.09 ± 1.13 | 12.25 ± 1.23  10.94 ± 0.32 |
| Cheese (grams/day) (JN)^*^ | 0.01 ± 0.05  -0.12 ± 0.06 | -0.12 ± 0.04  0.01 ± 0.06 | -0.04 ± 0.07  -0.07 ± 0.03 |
| French Fries (grams/day) | 23.51 ± 1.2  20.89 ± 1.29 | 19.91 ± 0.94  24.48 ± 1.32 | 22.96 ± 1.55  21.43 ± 0.63 |
| French Fries (grams/day) (JN)^*^ | 0.09 ± 0.05 | -0.05 ± 0.04 | 0.04 ± 0.06 |
| Moderate Physical Activity (MET Minutes/Week) | 547.6 ± 39.18  487.9 ± 39.72 | 462.09 ± 25.67  573.41 ± 44.63 | 491.77 ± 50.05  543.73 ± 18.58 |
| Moderate Physical Activity (MET Minutes/Week) (JN)^*^ | 0.21 ± 0.05  0.23 ± 0.05 | 0.14 ± 0.04  0.29 ± 0.05 | 0.18 ± 0.06  0.26 ± 0.03 |
| Negative Stress Subscale Score^*^ | 15.41 ± 0.25  15.45 ± 0.24 | 15.82 ± 0.19  15.04 ± 0.24 | 15.19 ± 0.3  15.67 ± 0.12 |
| Nightly Sleep Duration (hours)^**^ | 7.13 ± 0.08  7.38 ± 0.08 | 7.25 ± 0.07  7.26 ± 0.09 | 7.17 ± 0.1  7.34 ± 0.04 |

Post-study data are presented as model-adjusted least squares means ± SE. JN indicates JN transformed variables. Models were fit using baseline-adjusted linear regression in the combined sample from two randomized controlled trials. Fixed effects included Group, Sex, and Ethnicity, along with all two- and three-way interaction terms. All models adjusted for the baseline value of the outcome, baseline BMI, study, and state. Missing outcome data were addressed using multiple imputation by chained equations with predictive mean matching. Analyses were conducted within each imputed dataset, and estimates were combined using Rubin’s rules. Joint tests represent omnibus tests of each model term (e.g., Group, Sex, Ethnicity, and interaction terms) derived from pooled model estimates using the emmeans framework. Pairwise comparisons were calculated from the pooled estimates, with p-values adjusted using the multivariate t method.

P-values correspond to transformed variables unless transformation was not required to meet model assumptions, in which case non-transformed values are reported. Non-transformed data are presented for interpretability only when transformations were applied for analysis.

Superscript letters indicate pairwise differences for overall Group, Sex, and Ethnicity effects, shown only when higher-order interactions are not present. See Supplementary Table 3 for P-values of all main and interaction effects. ** indicates Control vs. Intervention; ^*^ indicates Female vs. Male; ¶ indicates Hispanic vs. Non-Hispanic.

BL; baseline, JN; Johnson’s Normalization transformation

**Supplemental Table 6. Model-Adjusted Significant Overall Group, Sex and Race Effects for Post-Study Outcomes in Young College Adults**

| **Outcome** | **Group:**  **Control**  **vs. Intervention** | **Sex:**  **Female**  **vs.**  **Male** | **Race:**  **Black**  **vs.**  **White**  **vs.**  **Other** |
| --- | --- | --- | --- |
| Waist Circumference (cm)^*^ | 81.29 ± 0.21   81.4 ± 0.28 | 81 ± 0.25  0.07 ± 0.01 | 0.08 ± 0.02  0.05 ± 0.01  0.05 ± 0.01 |
| NCIFV Score | 2.86 ± 0.12   3.39 ± 0.18 | 3.12 ± 0.15  3.12 ± 0.14 | 3.08 ± 0.28  3.08 ± 0.12  3.21 ± 0.16 |
| NCIFV Score (JN)^**^ | -0.18 ± 0.05   0.09 ± 0.05 | -0.09 ± 0.04   0 ± 0.05 | -0.04 ± 0.07  -0.04 ± 0.03  -0.06 ± 0.05 |
| Fruit and Fruit Juice (cups/day) | 1.08 ± 0.06  1.34 ± 0.06 | 1.14 ± 0.05   1.27 ± 0.07 | 1.18 ± 0.1  1.21 ± 0.04  1.24 ± 0.07 |
| Fruit and Fruit Juice (cups/day) (JN)^**^ | -0.18 ± 0.04   0.02 ± 0.05 | -0.13 ± 0.03  -0.02 ± 0.05 | -0.1 ± 0.07  -0.03 ± 0.03  -0.1 ± 0.05 |
| Fruit (cups/day) | 0.58 ± 0.04  0.75 ± 0.04 | -0.04 ± 0.04  -0.05 ± 0.05 | -0.02 ± 0.07  -0.03 ± 0.03  -0.09 ± 0.05 |
| Fruit (cups/day) (JN)^**^ | -0.16 ± 0.04  0.07 ± 0.05 | -0.04 ± 0.04  -0.05 ± 0.05 | -0.02 ± 0.07  -0.03 ± 0.03  -0.09 ± 0.05 |
| All Vegetables (cups/day) | 1.2 ± 0.06  1.4 ± 0.07 | 1.28 ± 0.06  1.31 ± 0.07 | 1.29 ± 0.1  1.21 ± 0.04  1.39 ± 0.08 |
| All Vegetables (cups/day) (JN)^**^ | -0.11 ± 0.05  0.11 ± 0.04 | -0.01 ± 0.04  0.01 ± 0.05 | -0.03 ± 0.08  -0.02 ± 0.03  0.04 ± 0.06 |
| All Vegetables (excluding lettuce and white potatoes, cups/day) | 0.54 ± 0.04  0.6 ± 0.05 | 0.58 ± 0.03  0.56 ± 0.05 | 0.53 ± 0.06  0.54 ± 0.03  0.64 ± 0.06 |
| All Vegetables (excluding lettuce and white potatoes, cups/day) (JN)^**^ | -0.07 ± 0.04  0.1 ± 0.04 | 0.03 ± 0.04  0 ± 0.05 | -0.01 ± 0.07  0.04 ± 0.03  0.01 ± 0.06 |
| Lettuce (cups/day) | 0.54 ± 0.04  0.7 ± 0.05 | 0.61 ± 0.03  0.63 ± 0.05 | 0.59 ± 0.07  0.57 ± 0.03  0.69 ± 0.05 |
| Lettuce (cups/day) (JN)^**^ | -0.12 ± 0.04  0.08 ± 0.04 | -0.04 ± 0.03  0 ± 0.05 | -0.07 ± 0.07  -0.02 ± 0.03  0.02 ± 0.05 |
| Cooked Dried Beans (cups/day) | 0.08 ± 0.01   0.1 ± 0.02 | 0.08 ± 0.01  0.11 ± 0.02 | 0.09 ± 0.02  0.09 ± 0.02  0.1 ± 0.01 |
| Cooked Dried Beans (cups/day) (JN)^*^ | -0.06 ± 0.05  0.02 ± 0.05 | -0.09 ± 0.04  0.05 ± 0.05 | -0.03 ± 0.07  -0.03 ± 0.04  -0.01 ± 0.06 |
| Tomato Sauce (cups/day) | 0.11 ± 0.01   0.13 ± 0.01 | 0.09 ± 0.01   0.14 ± 0.01 | 0.13 ± 0.02  0.11 ± 0.01  0.11 ± 0.01 |
| Tomato Sauce (cups/day) (JN)^*^ | -0.1 ± 0.05  -0.06 ± 0.05 | -0.16 ± 0.04  0 ± 0.06 | -0.06 ± 0.08  -0.06 ± 0.03  -0.12 ± 0.06 |
| Hot Dogs (grams/day) | 11.88 ± 1.1   13.23 ± 1.15 | 7.4 ± 0.7  17.71 ± 1.40 | 13.75 ± 1.55  9.48 ± 0.88  14.43 ± 1.41 |
| Hot Dogs (grams/day) (JN) ^¶, *^ | -0.02 ± 0.04  0.02 ± 0.04 | -0.18 ± 0.03  0.18 ± 0.05 | 0.09 ± 0.07  -0.11 ± 0.03  0.02 ± 0.06 |
| Cheese (grams/day) ^‡^ | 10.57 ± 0.49   10.68 ± 0.57 | 9.67 ± 0.42  11.58 ± 0.60 | 10.7 ± 0.83  11.69 ± 0.52  9.49 ± 0.58 |
| IPAQ Score^*^ | 2558.04 ± 90.08  265.89 ± 99.46 | 2391.14 ± 73.31  2823.79 ± 104.12 | 2454.25 ± 132.95  2753.19 ± 104.25  2614.96 ± 104.81 |
| Vigorous Physical Activity (MET minutes/week)^*^ | 1205.19 ± 61.64  1258.57 ± 68.92 | 1067.3 ± 48.28  1396.46 ± 79.24 | 1228.79 ± 94.63  1326.2 ± 81.14  1140.65 ± 70.94 |
| Moderate Physical Activity (MET Minutes/Week) | 505.74 ± 30.25   505.74 ± 30.25 | 438.84 ± 22.71  572.53 ± 32.69 | 442.39 ± 44.54  555.71 ± 20.7  518.95 ± 34.88 |
| Moderate Physical Activity (MET Minutes/Week) (JN) ^¶, *^ | 0.15 ± 0.04  0.25 ± 0.04 | 0.09 ± 0.03  0.3 ± 0.04 | 0.1 ± 0.06  0.29 ± 0.03  0.2 ± 0.05 |
| Total Stress Score^*^ | 25.14 ± 0.31  24.89 ± 0.37 | 25.59 ± 0.24  24.44 ± 0.43 | 25.53 ± 0.51  24.76 ± 0.43  24.76 ± 0.35 |
| Positive Stress Subscale Score^*^ | 18.57 ± 0.18  18.7 ± 0.19 | 18.39 ± 0.14   18.88 ± 0.20 | 18.41 ± 0.3  18.91 ± 0.12  18.58 ± 0.21 |
| Negative Stress Subscale Score^*^ | 15.64 ± 0.19   15.54 ± 0.22 | 16.09 ± 0.16  15.1 ± 0.23 | 15.92 ± 0.3  15.58 ± 0.2  15.28 ± 0.23 |
| Nightly Sleep Duration (hours)^**^ | 7.04 ± 0.07  7.43 0.1 | 7.23 ± 0.08  7.24 ± 0.08 | 7.15 ± 0.11  7.37 ± 0.07  7.19 ± 0.12 |

Post-study data are presented as model-adjusted least squares means ± SE. JN indicates JN transformed variables. Models were fit using baseline-adjusted linear regression in the combined sample from two randomized controlled trials. Fixed effects included Group, Sex, and Race, along with all two- and three-way interaction terms. All models adjusted for the baseline value of the outcome, baseline BMI, study, and state. Missing outcome data were addressed using multiple imputation by chained equations with predictive mean matching. Analyses were conducted within each imputed dataset, and estimates were combined using Rubin’s rules. Joint tests represent omnibus tests of each model term (e.g., Group, Sex, Race, and interaction terms) derived from pooled model estimates using the emmeans framework. Pairwise comparisons were calculated from the pooled estimates, with p-values adjusted using the multivariate t method.

P-values correspond to transformed variables unless transformation was not required to meet model assumptions, in which case non-transformed values are reported. Non-transformed data are presented for interpretability only when transformations were applied for analysis.

Superscript letters indicate pairwise differences for overall Group, Sex, and Race effects, shown only when higher-order interactions are not present. See Supplementary Table 4 for P-values of all main and interaction effects. ** indicates Control vs. Intervention; ^*^ indicates Female vs. Male; ¶ indicates Black vs. White; ‖ indicates Black vs. Other; ‡ indicates White vs. Other.

JN; Johnson’s Normalization transformation
